# Persistent symptoms and clinical findings in adults with post-acute sequelae of COVID-19/post-COVID-19 syndrome in the second year after acute infection: population-based, nested case-control study

**DOI:** 10.1101/2024.05.22.24307659

**Authors:** Raphael S. Peter, Alexandra Nieters, Siri Göpel, Uta Merle, Jürgen M. Steinacker, Peter Deibert, Birgit Friedmann-Bette, Andreas Niess, Barbara Müller, Claudia Schilling, Gunnar Erz, Roland Giesen, Veronika Götz, Karsten Keller, Philipp Maier, Lynn Matits, Sylvia Parthé, Martin Rehm, Jana Schellenberg, Ulrike Schempf, Mengyu Zhu, Hans-Georg Kräusslich, Dietrich Rothenbacher, Winfried V. Kern the EPILOC Phase 2 Study Group

## Abstract

**Objective:** To assess risk factors for persistence vs improvement and to describe clinical characteristics and diagnostic evaluation of subjects with post-acute sequelae of COVID-19/post-COVID-19 syndrome (PCS) persisting for more than one year.

**Design:** Nested population-based case-control study.

**Setting:** Comprehensive outpatient assessment, including neurocognitive, cardiopulmonary exercise, and laboratory testing in four university health centres in southwestern Germany (2022).

**Participants:** PCS cases aged 18 to 65 years with (n=982) and age and sex-matched controls without PCS (n=576) according to an earlier population-based questionnaire study (six to 12 months after acute infection, phase 1) consenting to provide follow-up information and to undergo clinical diagnostic assessment (phase 2, another 8.5 months [median] after phase 1).

**Main outcome measures:** Relative frequencies of symptoms and health problems and distribution of symptom scores and diagnostic test results between persistent cases and controls. Additional analysis included predictors of changing case or control status over time with adjustments for potentially confounding variables.

**Results:** At the time of clinical examination (phase 2), 67.6% of the initial cases (phase 1) remained cases, whereas 78.5% of the controls continued to report no health problems related to PCS. In adjusted analyses, predictors of improvement among cases were mild acute index infection, previous full-time employment, educational status, and no specialist consultation and not attending a rehabilitation programme. Among controls, predictors of new symptoms or worsening with PCS development were an intercurrent secondary SARS-CoV-2 infection and educational status. At phase 2, persistent cases were less frequently never smokers, had higher values for BMI and body fat, and had lower educational status than controls. Fatigue/exhaustion, neurocognitive disturbance, chest symptoms/breathlessness and anxiety/depression/sleep problems remained the predominant symptom clusters, and exercise intolerance with post-exertional malaise for >14 h (PEM) and symptoms compatible with ME/CFS (according to Canadian consensus criteria) were reported by 35.6% and 11.6% of persistent cases, respectively. In adjusted analyses, significant differences between persistent cases and stable controls (at phase 2) were observed for neurocognitive test performances, scores for perceived stress and subjective cognitive disturbances, symptoms indicating dysautonomia, depression and anxiety, sleep quality, fatigue, and quality of life. In persistent cases, handgrip strength, maximal oxygen consumption, and ventilator efficiency were significantly reduced. However, there were no differences in measures of systolic and diastolic cardiac function, in the level of pro-BNP blood levels or other laboratory measurements (including complement activity, serological markers of EBV reactivation, inflammatory and coagulation markers, cortisol, ACTH and DHEA-S serum levels). Screening for viral persistence (based on PCR in stool samples and SARS-CoV-2 spike antigen levels in plasma in a subgroup of the cases) was negative. Sensitivity analyses (pre-existing illness/comorbidity, obesity, PEM, medical care of the index acute infection) revealed similar findings and showed that persistent cases with PEM reported more pain symptoms and had worse results in almost all tests.

**Conclusions:** This nested population-based case-control study demonstrates that the majority of PCS cases do not recover in the second year of their illness, with patterns of reported symptoms remaining essentially similar, nonspecific and dominated by fatigue, exercise intolerance and cognitive complaints. We found objective signs of cognitive deficits and reduced exercise capacity likely to be unrelated to primary cardiac or pulmonary dysfunction in some of the cases, but there was no major pathology in laboratory investigations. A history of PEM >14 h which was associated with more severe symptoms as well as with more objective signs of disease may be a pragmatic means to stratify cases for disease severity.

**What is already known on this topic:** Self-reported health problems following SARS-CoV-2 infection have commonly been described and may persist for months. They typically include relatively non-specific complaints such as fatigue, exertional dyspnoea, concentration or memory disturbance and sleep problems. The incidence of this post-COVID-19 syndrome (PCS) is varying and associated with sociodemographic variables, pre-existing disease and comorbidities, the severity of the acute SARS-CoV-2 index infection, and some other factors. The long-term prognosis is unknown and may differ for different symptoms or symptom clusters. Evidence of measurable single or multiple organ dysfunction and pathology and their correlation with self-reported symptoms in patients with non-recovery from PCS for more than a year have not been well described.

**What this study adds:** The study describes the severity of the index infection, lower educational status, no previous full-time employment, and (need for) specialist consultation or a rehabilitation programme (the latter probably due to reverse causation) as factors for non-recovery from PCS, and found no major changes in symptom clusters among PCS cases persisting for more than a year. After a comprehensive medical evaluation of cases and controls and adjusted analyses, objective signs of organ dysfunction and pathology among persistent PCS cases correlated with self-reported symptoms, were detected more often among cases with longer lasting post-exertional malaise, and included both reduced physical exercise capacity (diminished handgrip strength, maximal oxygen consumption and ventilatory efficiency), and reduced cognitive test performances while there were no differences in the results of multiple laboratory investigations after adjustment for possible confounders.

## Introduction

The severe acute respiratory syndrome coronavirus 2 (SARS-CoV-2) pandemic resulted in over 750 million confirmed cases worldwide.[1] Besides morbidity and mortality in the acute phase of the infection, considerable post-acute health problems and sequelae are reported.[2–5] The WHO defined post-coronavirus disease 2019 [COVID-19] condition as the continuation or development of new symptoms after acute SARS-CoV-2 infection, lasting for at least two months, and being unexplained by an alternative diagnosis.[6] Slightly different definitions and alternative wording (such as long COVID-19 [LC]), post-acute sequelae of SARS-CoV-2 infection [PASC], or post-COVID-19 syndrome [PCS]) have been used[7,8] and are in part relevant for the widely differing prevalence estimates in previous studies.[9] Furthermore, previous prevalence estimates may have been biased since many of the early studies focused on hospitalized or healthcare-seeking patients only,[10–12] although most COVID-19 patients do not require medical treatment for the acute infection. Further limitations have been the difficulty of including an uninfected control group to estimate background prevalences of symptoms. In fact, many studies have assessed PCS prevalence and trajectories by using various questionnaires asking for self-reported health problems. Although many of the symptoms may impact everyday functioning, health-related quality of life and work ability,[3,4] they lack specificity (i.e. they can have many other causes and overlap with other conditions), are usually not well evaluable in claims data studies and have often not been validated through systematic protocol-prespecified diagnostic studies.

More recently, several diagnostic studies have been able to confirm some impaired neurocognitive functions[13–17] in PCS patients, while the results for cardiac and pulmonary function tests have been variable and less consistent.[18,19] Laboratory studies have suggested a number of altered blood biomarkers (such as various cytokines/chemokines, immune cell markers, plasma metabolites and cortisol) with potential pathophysiologic and diagnostic relevance in PCS patients.[20–23] Many of the clinical or laboratory diagnostic studies, however, were small, lacked appropriate controls, adjustments (e.g. for age and sex, smoking and body composition, educational or socioeconomic status, severity of the acute infection and pre-existing or concomitant disease), or showed only subtle changes compared to controls. Higher body mass index (BMI), for example, has been predictive for persisting dyspnoea in COVID-19 patients.[24] Obesity has been reported as a risk factor for PCS,[10,25,26] and mechanistic evidence of why obesity could make people more susceptible to PCS has been provided.[27] Outside the COVID-19 context, BMI in association with sex has been found to be a major confounder in studies[28] of proinflammatory markers, and obesity has also been associated with cognitive dysfunction.[29] Cognitive dysfunction, interestingly, has been measurable after COVID-19 in subjects who were asymptomatic or had no more symptoms than age- and sex-matched uninfected controls. [30,31] Symptom-based phenotypic stratification of PCS, although attractive and intriguing, thus, may be misleading in diagnostic studies if not evaluated against adequate controls and adjusted for potential confounders.

The aim of this study was to medically validate PCS cases that were defined as such in our previous population-based study of SARS-CoV-2 infected adults (6 to 12 months after infection) based on self-reported new symptoms with moderate to severe impairment in daily life plus either impaired general health or work ability.[32] From this population, we invited a number of PCS cases and controls (the latter being asymptomatic and reporting complete recovery from SARS-CoV-2 infection) to undergo a comprehensive outpatient medical examination and clinical evaluation, including standardized and validated questionnaires, neurocognitive and cardiopulmonary testing and laboratory investigations. We hypothesized that roughly half of the cases following the invitation would be persistent cases at the time of medical examination and expected that our clinical evaluation of persistent cases would result in an appreciable proportion of cases with measurable organ dysfunction and pathology and would show significant differences in at least one of the medical tests compared to stable controls. We were also interested in markers and risk factors for more severe disease and its possible underlying pathophysiology.

## Materials and methods

### Study design and selection of participants

This study was a prospective, multicentre, observational, nested case-control study. Subjects with (cases) and without PCS (controls) were recruited from the EPILOC (*<u>Epi</u>*demiology of *<u>Lo</u>*ng *<u>C</u>*ovid) phase 1 non-interventional, population-based questionnaire study that included subjects aged 18 to 65 years who had tested positive for SARS-CoV-2 by PCR between October 1^st^, 2020 and April 1^st^, 2021, and whose infection had been notified (compulsory according to the German Infection Protection Act) to the responsible local public health authority (in four administratively and geographically defined regions in the Federal State of Baden-Württemberg in southwestern Germany). Briefly, the study[32] (registered with “Deutsches Register Klinischer Studien”, DRKS 00027012) that was conducted six to 12 months after acute infection categorized 28.5% of the 11,710 evaluable respondents as suffering from PCS (cases), whereas 38% of the respondents were considered as (PCS-free) controls.

The PCS case definition used was “general health or working capacity recovered to a level no more than 80% (compared to pre-COVID-19), and any new symptom (a list of 30 symptoms was provided, three additional symptoms could be added) of moderate to strong degree regarding impairment in daily life and not already present before the acute infection (excluding vomiting, nausea, stomach ache, diarrhoea, chills, fever)”. Subjects who had recovered to >80% (of general health and work ability perceived in the time before acute infection) and reported no new symptoms of grade moderate to strong qualified as controls.

From the phase 1 PCS cases and controls, we invited participants into the phase 2 nested case-control study. A total of 982 cases and 576 frequency-matched age- and sex-matched controls followed the invitation and underwent a comprehensive clinical evaluation at one of the four study sites (**supplementary figure S1**). The unequal sampling ratio was based on the assumption that a significant number of phase 1 cases might have had recovered until presentation in phase 2, while we expected that only a small number of controls might have developed new symptoms compatible with PCS at the time of the clinical evaluation in phase 2. The study was registered with “Deutsches Register Klinischer Studien” (DRKS 00027362). All participants provided written informed consent, and the ethics committees of the respective universities approved the study.

### Data sources and measurements

Besides the information collected during the phase 1 study (see ref. [32]), we again used data from a number of standardized questionnaires that included sociodemographic characteristics, lifestyle factors, SARS-CoV-2 vaccines received, medical history and current symptoms. The symptom questionnaire contained the same items as in phase 1 and asked for medical treatment of current symptoms, for the grade to which each symptom impaired daily life and activities (“how much do you feel impaired by this at the moment?”) using a 4-point Likert scale (none, light, moderate, or strong) and for the degree of general health and working capacity regained (compared with the time before the index infection). We evaluated individual symptoms, but also symptom clusters composed of highly interrelated individual symptoms as defined earlier after analysis of the phase 1 study results.[32] Based on this information, we defined participants either as persistent (or improved) PCS case or as stably recovered control(dubbed “stable control”) or “worsened control”, using the same definition as in phase 1.

<u>Clinical assessments</u>: Apart from taking the medical history, the study physician completed a modified Medical Research Council Dyspnoea Scale (mMRC), asked for post-exertional malaise and its duration,[33] and clarified questions and responses to the questionnaires. The participants underwent a complete physical examination, including measurements of height, weight, heart rate (HR) at rest, and blood pressure.

The maximal grip strength was recorded after three measurements of both hands with a digital hand dynamometer. Whole body composition was measured using a multifrequency bioelectrical impedance analysis device and expressed as % body fat. Methodological details are included in **supplementary text S1**.

<u>Validated questionnaires</u>: Study participants were asked to fill validated questionnaires on sleep quality (Pittsburgh Sleep Quality Index [PSQI], Insomnia Severity Index [ISI], Epworth Sleepiness Scale [ESS]), fatigue (Chalder Fatigue Scale [CFQ-11]), health-related quality of life (Short Form-12 Health Survey [SF-12]), assessing both physical and mental components), symptoms of depression (Patient Health Questionnaire 9 [PHQ-9]), anxiety (Generalised Anxiety Disorder 7 [GAD-7]), perceived stress (10-item Perceived Stress Scale [PSS-10]), subjective cognition (“Fragebogen zur geistigen Leistungsfähigkeit” [FLei]), and dysautonomia symptoms (Composite Autonomic Symptom Score 31 [COMPASS-31]). More details and references are given in **supplementary text S1**).

<u>Neurocognitive tests</u>: All subjects were asked to undergo neuropsychological tests administered by trained clinical staff. The test battery included the Montreal Cognitive Assessment (MoCA), the Trail Making Test part B (TMT-B), and the Symbol Digit Modalities Test (SDMT) (**supplementary text S1**).

<u>Cardiopulmonary function tests</u>: We recorded resting 12-lead electrocardiograms (ECG) and pulse oximeter measurements of peripheral oxygen saturation (SpO_2_). Resting echocardiograms were performed according to current guidelines, with determination of the left ventricular volume and ejection fraction (LV-EF), the ratio between early mitral inflow and mitral annular early diastolic velocities (LV-E/e’), the ratio of maximal early to late diastolic transmitral flow velocity (LV-E/A), and grading of diastolic dysfunction (**supplementary text S2**).

Participants underwent cardiopulmonary exercise testing (CPET) using a ramp protocol on the cycle ergometer. Before CPET, spirometry was conducted to assess lung function with recording of the forced expiratory volume in one second (FEV1), and the forced vital capacity (FVC). During CPET, blood pressure, SpO_2_, and ECG with HR were monitored. We evaluated the following CPET parameters: HR, oxygen uptake (VO_2max_), breathing reserve (BR), respiratory exchange ratio (RER), and the slope of minute ventilation to carbon dioxide production (VE/VCO_2_ slope). More details are included in **supplementary text S2**.

<u>Laboratory investigations</u>: Routine laboratory investigations included a rapid chromatographic immunoassay (for SARS-CoV-2 antigen in nasopharyngeal samples), blood cell counts, coagulation, clinical chemistry, levels of C-reactive protein (CRP), thyroid stimulating hormone (TSH), glycated haemoglobin (HbA1c), N-terminal pro-brain natriuretic peptide (proBNP), classical pathway complement haemolytic activity (CH50) (determined for participants at two centres), antibodies against CMV, SARS-CoV-2 nucleocapsid (N) protein and the S1 receptor binding domain of the viral spike glycoprotein, and others (see **supplementary text S3** for analytes and methods). Cortisol, ACTH and DHEA-S levels in frozen morning blood samples were measured centrally using standard methods (see **supplementary text S3** for details). Additional laboratory investigations in our central virology laboratory included the measurement of antibodies to Epstein-Barr virus (EBV) antigens, of spike antigen in serum (in a subgroup of persistent cases and controls), and SARS-CoV-2 RNA by RT-PCR in faecal samples (see **supplementary text S3** for detailed methodologies).

### Statistical methods

Participant characteristics were analysed descriptively. Predictors of case-control status change from phase 1 to phase 2 were evaluated using logistic regression. Regression models were run separately for phase 1 cases and phase 1 controls and mutually adjusted odds ratios were calculated for improvement in cases (no longer fulfilling the case definition) and worsening in controls (no longer fulfilling the control definition).

Results of standardised questionnaires, neurocognitive tests, laboratory measurements, electrocardiographic, echocardiographic, and spiroergometric parameters were presented as least square means for persistent cases, improved cases, worsened controls and stable controls. Due to a high correlation between PSQI, ISI, and ESS, we present only the results for the PSQI instrument.

We used analysis of covariance with adjustment for sex-age class combinations and university entrance qualification. Additional adjustments were made as indicated. Geometric instead of natural means are reported where appropriate. The area under the receiver operating characteristic curve (AUC) for discrimination of persistent cases versus stable controls (excluding improved cases and worsened controls), based on logistic regression, is also reported. Statistical procedures were performed with the SAS statistical software package (release 9.4 SAS Institute Inc.) or R version 4.3.2.

### Patient and public involvement

This study was conducted in rapid response to the SARS-CoV-2 pandemic, a public health emergency of national and international concern. Neither patients nor the public were directly involved in the design, conduct, or reporting of this research. We were aware from engagement of and discussion with patient support groups that further information on the medium- and long-term prognosis of PCS and a comparison with ME/CFS were desired.

## Results

### Baseline characteristics of the study participants

The study included 982 participants who were phase 1 cases and 576 age and sex-matched phase 1 controls. As shown in **supplementary table S1**, the sex and age distributions were (as expected by design) similar in cases and controls. Most (circa 65%) participants were female, and the mean age was 48 years. The mean time between phases 1 and 2 was 9.1 months in cases (range 3.0 to 14.2 months) and 8.4 months in controls (range 2.9 to 14.0 months), respectively. A similar proportion of cases versus controls experienced a secondary SARS-CoV-2 infection (23%) and almost all had been vaccinated against SARS-CoV-2 once or more times before phase 2 (**supplementary table S1**).

Differences between cases and controls already known from the analysis of phase 1 data included the proportion of obese participants, smokers, pre-existing diseases, medical care (outpatient or inpatient versus none) for the earlier index acute SARS-CoV-2 infection (each higher among cases), and educational level (fewer cases with university entrance qualification). Healthcare utilization in the last 6 months prior to phase 2 examination (in particular regarding specialist physician consultation) and attending a rehabilitation programme were also much more frequent among cases. **Supplementary figure S2** describes the probability of participation in cases and controls by selected baseline characteristics.

### Risk for PCS persistence

Roughly two-thirds (67.6%) of the 982 participants classified as phase 1 cases were considered persistent cases (according to our PCS case working definition) after the phase 2 clinical assessment. Most of the remaining phase 1 cases (30.1%) had improved until phase 2, but only very few (2.2%) were classified as complete clinical recovery (**figure 1**). Conversely, the majority (78.5%) of controls from phase 1 who participated in phase 2 were classified as stable controls, but almost one-fifth (18.9%) reported new symptoms (without fulfilling the PCS case definition), and 2.6% were classified as (new-onset) PCS cases (**figure 1**). **Supplementary figure S3** displays changes in the prevalence of the five main symptom clusters among the participating phase 1 cases as evaluated in phase 2. In the overall population, the net prevalence of all symptom clusters, except anxiety, depression or sleep disorder decreased, most prominent for smell and taste disorders (**supplementary figure S3**).

**Figure 1.**
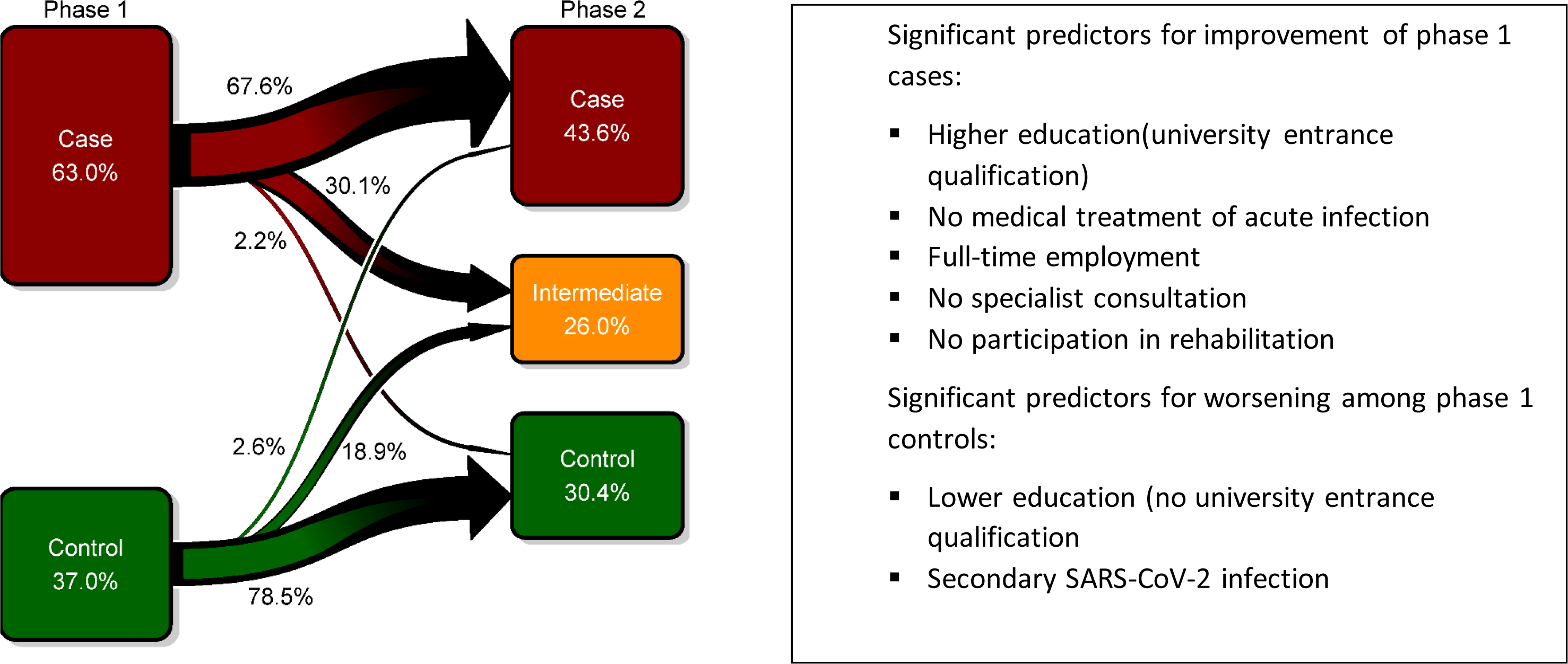
Change in case/control status of study participants (N=1558) between initial questionnaire survey (phase 1) and clinical examination (phase 2). The time from PCR-confirmed acute SARS-CoV-2 infection to phase 1 was 8.7 months (median), the time from phase 1 participation until clinical examination in phase 2 was 8.5 months (median), and the median time between acute infection and phase 2 was 17.2 months, ranging from 9.2 to 24.4 months. Significant predictors for improvement of phase 1 cases and for worsening among phase 1 controls were assessed after calculation of ORs with mutual adjustment for the following variables: sex, age, university entrance qualification, marital status, medical treatment of acute infection, obesity (BMI ≥ 30kg/m²), full-time employment (phase 1), time between phase 1 and phase 2 (per month), secondary SARS-CoV-2 infection since phase 1, two or more vaccine shots, (any) specialist consultation in the last six months, participation in a post-COVID-rehabilitation program (see supplementary table 2).

As summarized in **figure 1** (and detailed in **supplementary table S2**), predictors of improvement (either to intermediate or control status) among cases in an adjusted analysis were educational status (university entrance qualification), full-time employment (at phase 1), no medical care/treatment of the acute index infection (as a proxy for milder acute infection), and no (need for) specialist consultation within the last six months or participation in a post-COVID-19 rehabilitation program (the latter two probably a result of reverse causation). For controls, the odds of worsening until phase 2 were higher with lower educational status and after a secondary SARS-CoV-2 infection since phase 1. SARS-CoV-2 vaccination had no measurable association with improvement in cases or worsening in controls. Also, age, sex, or the time between phase 1 and phase 2 was not statistically significantly associated with case-control status changes (**supplementary table S2**).

### Clinical evaluation of persistent PCS cases

In a comparison of the characteristics of persistent cases with those of improved cases, worsened controls and stable controls (**table 1**), we found differences in educational status, smoking, BMI (as well as obesity prevalence and body fat), medical care/treatment of the acute SARS-CoV-2 index infection and prevalence of comorbidities. The proportion of participants with obesity was highest in persistent cases (30.2% compared with 12.4% in stable controls), and many more stable controls than persistent cases have had no medical care for their acute index infection, had obtained university entrance qualification and were never smokers ( **table 1**). We found a much higher current use of medication in persistent cases versus stable controls across all anatomical-therapeutic-chemical (ATC) groups (**supplementary table S3**). The three drug classes with the largest ratio between persistent cases and stable controls were ATC N06 (including antidepressants), drugs against peptic ulcer disease and reflux (ATC A02B), and beta blockers (ATC C07).

**Table 1.** Characteristics of the phase 2 study participants by case-control status.

|  | Persistent cases |  | Cases improved |  | Controls worsened |  | Stable controls |  |
| --- | --- | --- | --- | --- | --- | --- | --- | --- |
|  | N | Mean or frequency | N | Mean or frequency | N | Mean or frequency | N | Mean or frequency |
| Male, n (%) | 664 | 227 (34.2) | 318 | 122 (38.4) | 124 | 44 (35.5) | 452 | 153 (33.9) |
| Female, n (%) |  | 437 (65.8) |  | 196 (61.6) |  | 80 (64.5) |  | 299 (66.2) |
| Age at phase 1 (years), mean (SD) |  | 48.9 (12.1) |  | 46.3 (12.5) |  | 48.4 (11.9) |  | 48.5 (12.4) |
| Age class at phase 1 (years), n (%) |  |  |  |  |  |  |  |  |
| 18-29 | 664 | 74 (11.1) | 318 | 49 (15.4) | 124 | 14 (11.3) | 452 | 55 (12.2) |
| 30-39 |  | 76 (11.5) |  | 50 (15.7) |  | 14 (11.3) |  | 55 (12.2) |
| 40-49 |  | 128 (19.3) |  | 60 (18.9) |  | 27 (21.8) |  | 91 (20.1) |
| 50-59 |  | 267 (40.2) |  | 116 (36.5) |  | 49 (39.5) |  | 159 (35.2) |
| 60+ |  | 119 (17.9) |  | 43 (13.5) |  | 20 (16.1) |  | 92 (20.4) |
| University entrance qualification, n (%) | 664 | 257 (38.7) | 318 | 163 (51.3) | 124 | 60 (48.4) | 452 | 278 (61.5) |
| Full-time employment at phase 1, n (%) | 663 | 306 (46.2) | 318 | 194 (61.0) | 124 | 66 (53.2) | 451 | 223 (49.5) |
| Smoking status, n (%) |  |  |  |  |  |  |  |  |
| Current | 662 | 52 (7.9) | 317 | 20 (6.3) | 124 | 10 (8.1) | 452 | 17 (3.8) |
| Former |  | 205 (31.0) |  | 78 (24.6) |  | 36 (29.0) |  | 93 (20.6) |
| Never |  | 405 (61.2) |  | 219 (69.1) |  | 78 (62.9) |  | 342 (75.7) |
| BMI at phase 2 (kg/m<sup>2</sup>), mean (SD) | 662 | 28.0 (6.1) | 318 | 26.6 (5.5) | 124 | 26.1 (4.5) | 452 | 25.0 (4.5) |
| Obese (≥30 kg/m<sup>2</sup>), n (%) |  | 200 (30.2) |  | 64 (20.1) |  | 25 (20.2) |  | 56 (12.4) |
| Body fat (per cent), mean (SD) | 659 | 32.2 (10.6) | 316 | 29.3 (9.4) | 123 | 28.5 (9.0) | 452 | 27.4 (8.9) |
| >25% in men, >35% in women, n (%) |  | 344 (52.2) |  | 122 (38.6) |  | 45 (36.6) |  | 126 (27.9) |
| Treatment of acute SARS-CoV-2 infection, n (%) |  |  |  |  |  |  |  |  |
| No medical care | 655 | 341 (52.1) | 313 | 200 (63.9) | 123 | 108 (87.8) | 450 | 408 (90.7) |
| Outpatient care |  | 258 (39.4) |  | 92 (29.4) |  | 12 (9.8) |  | 37 (8.2) |
| Inpatient care (without ICU) |  | 45 (6.9) |  | 17 (5.4) |  | 3 (2.4) |  | 3 (0.7) |
| Intensive care |  | 11 (1.7) |  | 4 (1.3) |  | 0 (0.0) |  | 2 (0.4) |
| Comorbidities, n (%) |  |  |  |  |  |  |  |  |
| Cardiovascular disease | 664 | 29 (4.4) | 318 | 2 (0.6) | 124 | 1 (0.8) | 452 | 3 (0.7) |
| Chronic pulmonary disease |  | 62 (9.3) |  | 34 (10.7) |  | 6 (4.8) |  | 23 (5.1) |
| Diabetes mellitus |  | 33 (5.0) |  | 8 (2.5) |  | 2 (1.6) |  | 5 (1.1) |
| Cancer |  | 13 (2.0) |  | 3 (0.9) |  | 1 (0.8) |  | 4 (0.9) |

<u>Predominant symptoms, symptom clusters and symptom severity</u>: An analysis among persistent cases of the frequency of all reported symptoms with all degrees of impairment (**supplementary figure S4**) showed the predominance of individual complaints and symptoms that we summarize in the symptom clusters “fatigue”, “neurocognitive disturbance”, “chest symptoms”, “smell or taste disorder”, and “anxiety/depression/sleep disorder”. As shown in **supplementary figure S4**, there were some differences in individual symptom prevalence and severity between females and males (with females being more affected - similar to findings in phase 1), and several individual symptoms were scored comparatively low regarding their grade of daily life impairment (for example dizziness, paraesthesia, confusion and chest pain). Abdominal symptoms, fever and chills, and skin problems were rare, similar to what we found in phase 1.

We next displayed the distribution of (case-defining, i.e. moderate or severe) predominant symptoms and symptom clusters among persistent cases versus the other subgroups, together with the scoring results from corresponding validated questionnaires either as proportions at relevant cut-offs (**table 2**) or as adjusted average ratings (**figure 2**). As shown in **table 2**, fatigue, neurocognitive disturbance, and chest symptoms were among the predominant symptom clusters of persistent cases. We observed a large overlap of these three clusters among persistent cases, with a substantial proportion (26.8%) reporting moderate or severe symptoms in all three main symptom clusters (**supplementary figure S5**). The second largest overlap was the combination of fatigue and neurocognitive disturbance (prevalence, 20.1%). These three main symptom clusters comprised the vast majority (90.4%) of persistent cases.

**Figure 2.**
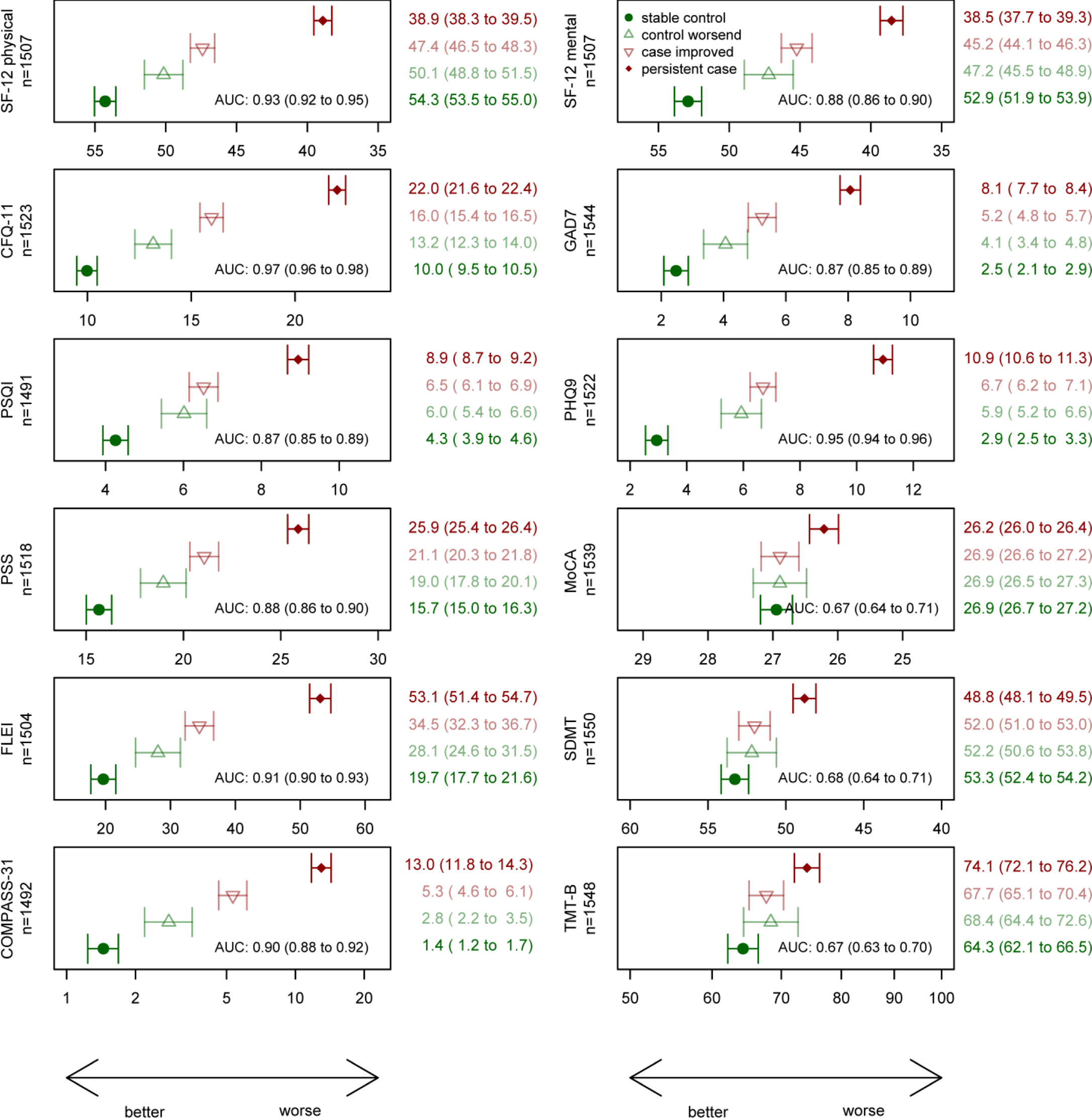
Means (geometric mean for COMPASS-31 and TMT-B) of self-reported health outcomes and neurocognitive tests (with 95%-CI) by case-control status at clinical examination in phase 2, adjusted for sex-age class combinations, study centre, and university entrance qualification. The reported area under the curve (AUC) for persistent cases vs. stable controls by the respective instrument also includes sex-age class combinations and university entrance qualification. The AUC for sex -age class combinations, study centre and university entrance qualification alone was 0.64. For comparability, the x-axis is scaled from mean -1 SD to mean +1 SD for all panels. MoCA: Montreal cognitive assessment scale (points); SDMT: Symbol Digit Modalities Test (number of correct symbols); TMT-B: Trail making test B (time in seconds).

**Table 2.**
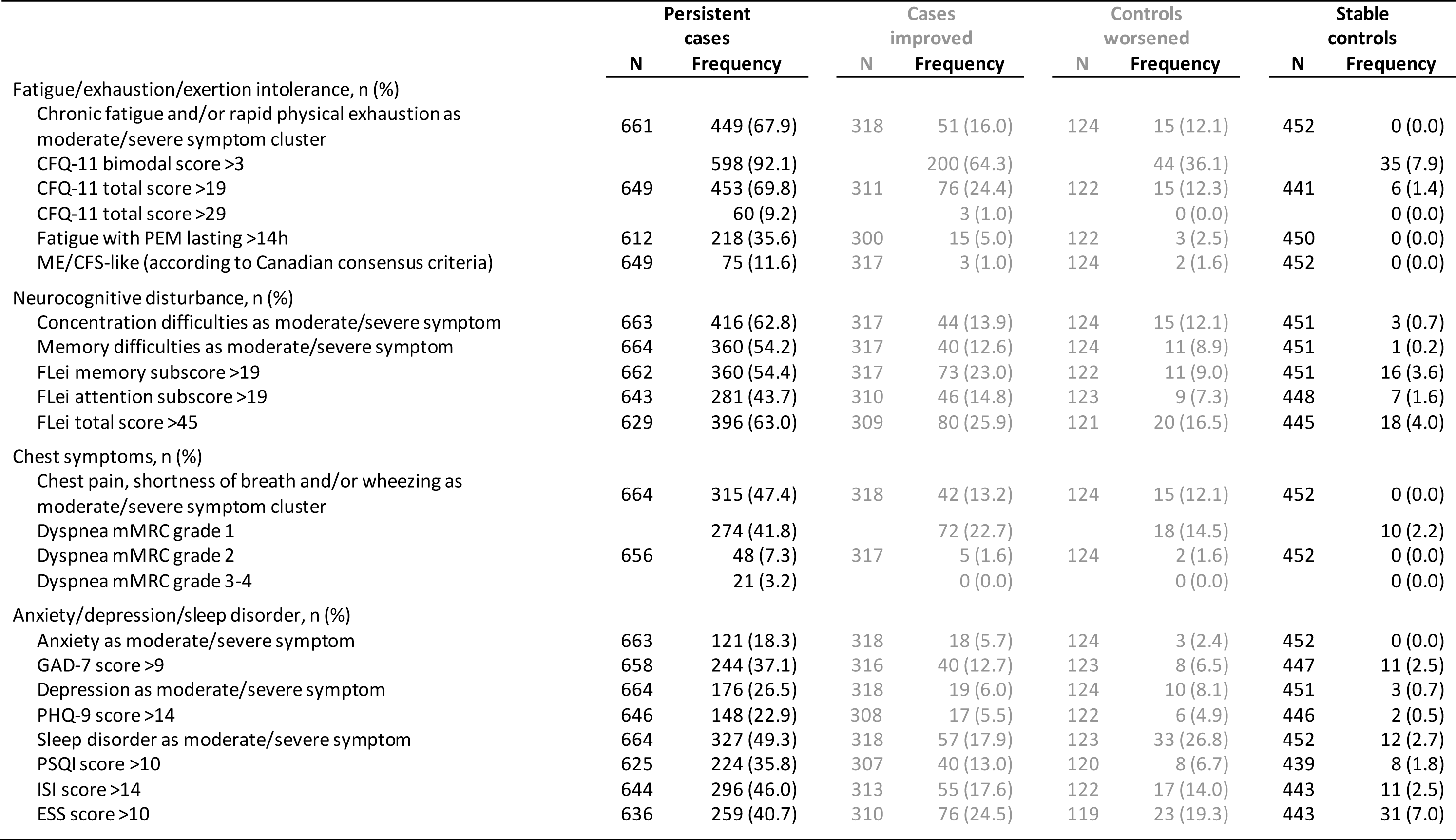
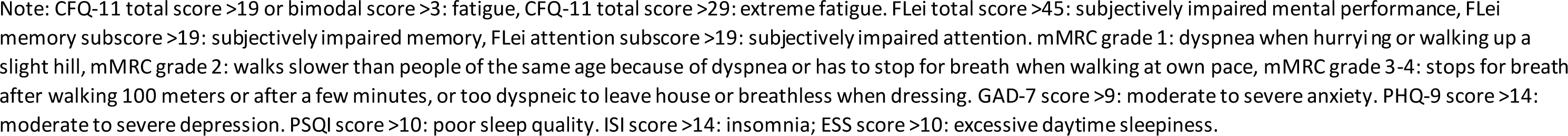
Prevalence of major symptom new clusters/symptoms and associated severity ratings according to validated questionnaires by ca se-control status at clinical examination in phase 2.

The frequency estimates for a given symptom or symptom cluster varied somehow with more detailed questioning or rating, allowing a more valid estimation of severity. Fatigue as the most prevalent self-reported symptom cluster (based on reporting chronic fatigue or rapid physical exhaustion of moderate or strong grade in the symptom questionnaire), for example, had a prevalence among persistent cases of 67.6%, while the prevalence assessed with the CFQ-11 scale at a bimodal score >3 (earlier defined as a “fatigue case”) or at a total score >19 was 92.1% and 69.8%, respectively. The prevalence of extreme fatigue (CFQ-11 total score >29) was relatively low among persistent cases (9.2%) (**table 2**).

We also assessed the prevalence of fatigue with PEM lasting >14 hours (35.6%) and of symptoms compatible with an ME/CFS-like condition (11.6%). Interestingly, the frequency of individual symptoms (of any degree) among PEM (lasting >14 hours) cases differed somehow from those persistent cases who had no PEM. Persistent cases with PEM had more symptoms than persistent cases without PEM. In particular, pain syndromes (chest pain, myalgia, joint pain, melalgia, headache), confusion and dizziness were more often reported by cases with PEM (apart from fatigue and exhaustion) (**supplementary figure S4**). PEM was highly prevalent (>50%) among persistent cases who reported symptoms from all three dominant clusters (fatigue, neurocognitive disturbances, chest symptoms) (**supplementary figure S6**).

Neurocognitive impairment remained the second most frequent symptom cluster (per symptom questionnaire) after fatigue in persistent cases, which correlated well with the FLei questionnaire results (**table 2**). Concentration difficulties were slightly more often reported than memory difficulties, and using a FLei memory subscore at a cut-off >19 confirmed this pattern. Dyspnea was most often non-severe when assessed with mMRC grading (**table 2**). The prevalence of mMRC grade 1 dyspnea among persistent cases was 41.8%, and dyspnea of grade 2 or more was seen in 10.5%. Symptoms of anxiety, depression, and sleep disorders (that had earlier been classified as a single cluster of highly interrelated symptoms) were also much more prevalent among persistent cases than among stable controls. Both sleep problems and depressive symptoms, as reported via the symptom questionnaire, interestingly, appeared somehow overrated when compared with the results of the validated questionnaires at conventional cut-offs (**table 2**), whereas anxiety as (moderate or severe) symptom appeared somehow underrated compared with the frequency of a GAD-7 score >9 (suggesting moderate to severe anxiety).

The average scores of CFQ-11, FLei, GAD-7, PHQ-9 and PSQI differed substantially and consistently between persistent cases and the other subgroups, and all these instruments discriminated persistent cases from stable controls very well, with the CFQ-11 having the highest AUC (>0.90) (**figure 2**).

<u>Symptoms of dysautonomia</u>: Based on earlier experience that autonomic nervous system dysfunction may be common among patients with long COVID-19 or with ME/CFS, we included the COMPASS-31 instrument as a screening questionnaire covering the history of orthostatic intolerance and other components of dysautonomia. As shown in **figure 2**, the average COMPASS-31 score among persistent cases was 13 compared with <2 among stable controls, and the proportion of persistent cases with a score >19 (suggesting moderate or severe dysautonomia) was 40.7%. Almost half of the persistent cases (49.7% compared with 7.5% of stable controls) indicated that they experienced weakness, dizziness, lightheadedness, or difficulty thinking after standing up from sitting or lying down, suggesting orthostatic problems.

<u>Perceived stress and health-related quality of life</u>: As a measure of stress and health-related quality of life, we used the PSS-10 instrument (scoring from 0 to 40) and the commonly used SF-12 questionnaire with its physical and mental component summary scores, assessing general health and well-being, including the impact of any illnesses or adverse condition on a broad range of functional domains. As shown in **figure 2**, all three scores discriminated well between persistent cases and stable controls and had similarly high AUCs >0.8, showing strong discriminative ability. The differences in the average scores between persistent versus improved cases and between stable versus worsening controls showed a similar pattern as the other instruments. A direct comparison of the current SF-12 results for both components in persistent cases with those obtained earlier in the same subjects (at phase 1) indicated no improvement in health-related quality of life in persistent cases (data not shown).

<u>Neurocognitive testing</u>: The results of the three neurocognitive tests are depicted in **figure 2**. In adjusted analysis, the mean MoCA score was significantly lower among persistent cases compared with the other groups, and the number of participants with a MoCA score below 26 (suggesting mild to moderate cognitive impairment) was 33.3% among persistent cases and 18.9% among stable controls, respectively. Similar patterns were seen with the two other tests, SDMT (assessing impaired attention, concentration and speed of information processing) and TMT-B (to screen executive dysfunction). Although the mean differences between persistent cases and stable controls were large, the discrimination in adjusted analysis between the two groups, however, was relatively poor for each test (AUCs 0.67 compared to 0.63 without neurocognitive testing). Further adjustment for CFQ-11 and PHQ-9 attenuated the association with MoCA to some degree, with differences for persistent cases versus stable controls losing statistical significance (p=0.0672). However, the additional adjustment had little effect on the association with SDMT and TMT-B (p=0.0086 and p=0.0008).

<u>Grip strength and cardiopulmonary function tests</u>: The mean maximal handgrip strength among persistent cases was 40.2 kg, significantly lower than that of stable controls (42.5 kg) (**figure 3**). As expected, grip strength was lower in women than men and associated inversely with body fat and BMI (data not shown). As depicted in **figure 3**, left ventricular function (including LV-EF, LV-E/e’, and LV-E/A) and pro-BNP blood levels were not different between the groups. We observed a higher prevalence of diastolic dysfunction grade 1 and 2 among persistent cases compared with controls (30.9% versus 21.9%) (**supplementary table S4**). The difference, however, was not statistically significant after adjustment for sex-age class combinations, study centre, university entrance qualification, BMI and smoking status. We also did not observe differences between the subgroups in the mean values for resting HR and BR (**figure 3**), respiratory rate and systolic and diastolic blood pressure (data not shown).

**Figure 3.**
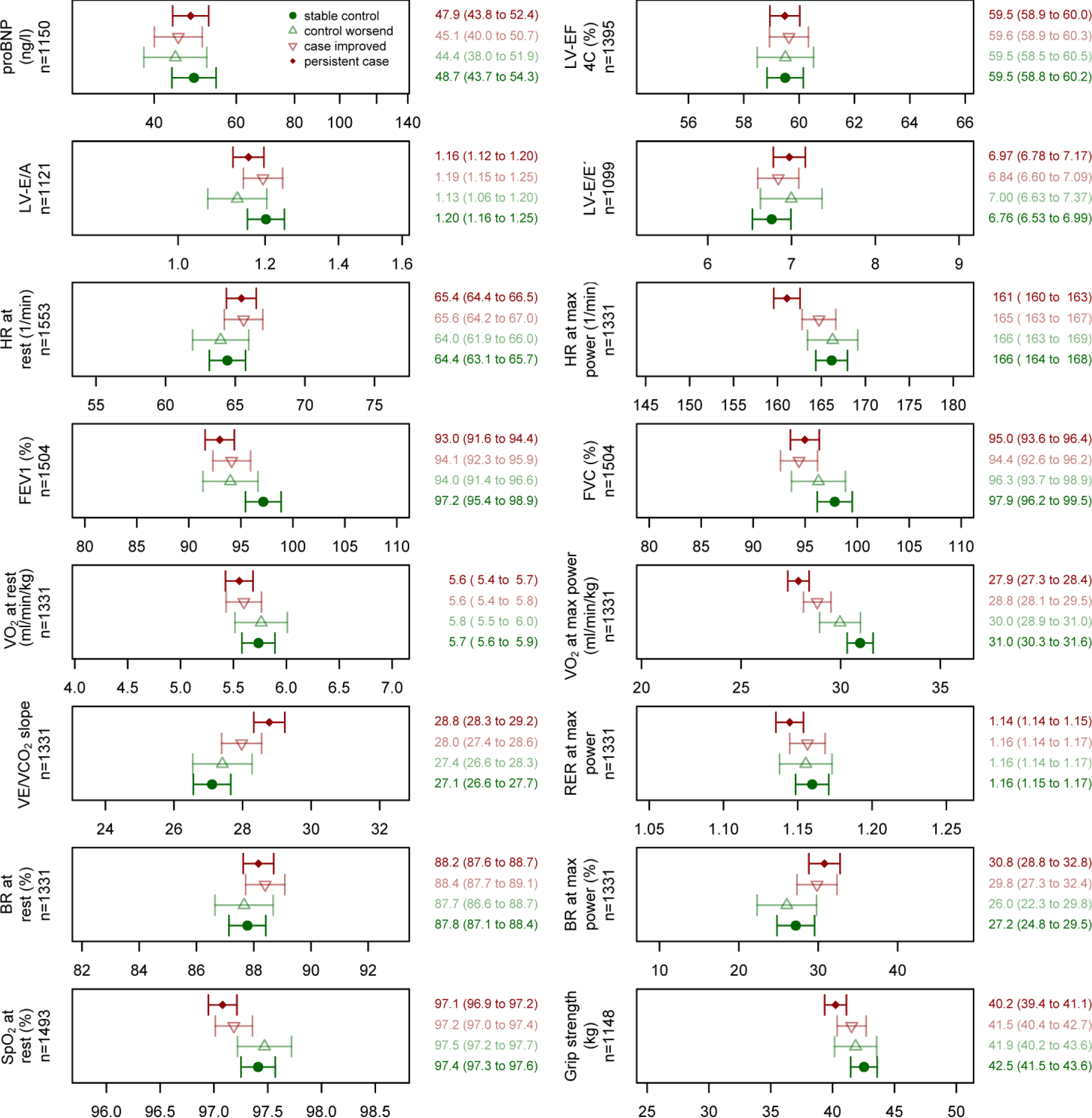
Cardiopulmonary function indicators and grip strength (means with 95%-CI) by case-control status at the clinical examination in phase 2, adjusted for sex-age class combinations, study centre, university entrance qualification, BMI, smoking status and use of beta blocking agents. Cardiopulmonary exercise testing could be completed in 1331 participants (87.2% of stable controls, 83.7% of persistent cases). For comparability, the x-axis is scaled from mean -1 SD to mean +1 SD for all panels.

Differences were observed for FEV1 and FVC, SpO2 at rest, and several CPET derived variables. Values for FEV1 (p<0.0001), FVC (p=0.0011), and SpO_2_ (p=0.0001) were lower among persistent cases (versus stable controls), but the differences were small, and the proportion of subjects with FEV1/FVC <0.70 was similar in persistent cases versus stable controls (10.3% versus 9.6%) (**supplementary table S4**).

In CPET, persistent cases achieved a lower maximal power with lower HR than the participants of the other subgroups, but RER values at the end of CPET were similar and well above 1.05, indicating exhaustion and attaining VO_2max_. Also, the median values of the Borg CR10 scale were similar for persistent cases and controls (data not shown). The most relevant and significant CPET differences between persistent cases and controls were observed for VE/VCO_2_ slope (higher values in persistent cases) and VO_2max_ (lower values in persistent cases) (**figure 3**). The proportion of persistent cases with VO_2max_ <85% of target value (suggesting reduced exercise capacity possibly due to deconditioning or peripheral muscle limitations) was significantly greater than that of stable controls in adjusted analyses (35.3% versus 8.4%) (**supplementary table S4**). Similarly, the differences in the proportion of participants with VO_2max_ below defined thresholds for males and females was substantial and highly significant between persistent cases and stable controls (**supplementary table S4**). Furthermore, we detected a significant difference in the mean VE/VCO_2_ slope between persistent cases and stable controls (28.8 versus 27.1) (**figure 3**), resulting in a higher proportion of persistent cases (versus stable controls) with values >30 (34.9% versus 18.5%) or >34 (13.5% versus 4.1%) (**supplementary table S4**).

We explored a possible overlap of objective signs of cognition deficits and reduced cardiorespiratory capacity within the persistent PCS case population ( **supplementary table S5**). The proportion of persistent PCS cases with MoCA ≤25 and SDMT <36 increased with increasing VE/VCO_2_ slope, and there were more cases with SDMT <36 in persistent cases with poor VO_2max_ (<85% predicted), but there were no such results for TMT-B, and persistent cases with differences in the Tiffenau test did not differ in their cognitive test performances (**supplementary table S5**).

<u>Laboratory investigations</u>: Besides pro-BNP (see above and **figure 3**), we measured complete blood counts, several blood levels including CRP, LSH, ferritin, liver and renal function and coagulation markers (D-dimer, von Willebrand factor [vWF] antigen and activity), TSH, cortisol, ACTH, DHEA-S, HbA1c, 25-hydroxy-vitamin D3, CH50, and others. After adjustment for sex-age class combinations, study centre, university entrance qualification, BMI and smoking status, we found no significant differences between persistent cases and stable controls in any of these laboratory investigations (**supplementary figures S7** and **S8**, **supplementary table S6**, **supplementary text S3**). Notably, there was a statistically significant association for CRP, HbA1c, and D-dimers before adjustment for BMI and smoking (data not shown).

We did not observe significant differences between persistent cases and stable controls in the prevalence of S1 and N SARS-CoV-2 antibodies (data not shown) or in the level of antibodies against SARS-CoV-2 S1 antigen (**supplementary figure S9**). Also, positivity rates for antibodies against CMV and several EBV antigens (VCA, EBNA, and EA-D) did not differ significantly between groups (**supplementary table S7**). The proportion of study participants with EBV serology indicative of reactivation was 13% (194 of 1,468 seropositive participants). However, we detected no elevated risk for EBV reactivation among persistent cases or controls reporting new symptoms between phases 1 and 2 (**supplementary table S7**). We additionally looked at EA-D and EBNA IgG antibody levels in participants with evidence for EBV reactivation but did not detect differences between persistent cases and stable controls with or without PEM (**supplementary figure S10**).

All participants were negative for SARS-CoV-2 antigen in oropharyngeal swabs by a rapid antigen assay at presentation. Using an ultrasensitive antigen ECL assay, we could not detect SARS-CoV-2 spike antigen in plasma samples from a subgroup of 100 persistent cases and 100 stable controls. Also, RT-PCR for SARS-CoV-2 RNA was negative in all tested stool samples from a similar subgroup of 156 persistent cases and 103 stable controls (see also **supplementary text S3**), allowing to state with a certainty of 95% that the true PCR positivity prevalence in persistent cases 17 months after infection is less or equal to 1.9%.

### Sensitivity analyses

The results of several sensitivity analyses (pre-existing illness/comorbidity, obesity, PEM, medical care of the index acute infection) are presented in supplementary figures. The general patterns persisted as described previously, and the differences in the validated questionnaire scores, in neurocognitive as well as in cardiopulmonary tests that were significant in the full analysis set, remained significant (**supplementary figures S11** to **S20**). The odds of finding abnormal neurocognitive and cardiopulmonary test results were higher for female participants with persistent PCS than for male participants with persistent PCS, but the differences were significant only for the TMT-B test (**supplementary figure S21**).

We also show that in the subpopulation of participants without preexisting diseases and comorbidity, the changes between phases 1 and 2 in the prevalence of main symptom clusters among cases were similar to those observed in the full analysis ( **supplementary figure S3**). When persistent cases were stratified according to PEM (lasting >14 hours), the burden of symptoms and complaints as reported and as assessed by validated questionnaires was much higher among persistent cases with versus those without PEM symptoms, including sleep problems, depression and anxiety, perceived stress and subjective cognition impairment, fatigue and dysautonomia (**supplementary figures S4** and **S17**). The analysis of neurocognitive testing also showed PEM to be associated with substantially worse results ( **supplementary figure S17**), particularly in the SDMT, which assesses cognitive processing speed. However, persistent cases without PEM still had significantly worse results than stable controls in all three tests. Persistent cases with PEM also showed reduced handgrip strength, lower oxygen saturation, lower peak heart rate, higher values for VE/VCO_2_ slope, and reduced VO_2max_ when compared with persistent cases without PEM (**supplementary figure S18**), and the proportion with VO_2max_ <85% of target value was high (41.0% versus 32.5% in persistent cases with versus without PEM [data not shown]). Several other variables of cardiopulmonary function differed between the two subgroups (**supplementary figure S18**), although some showed only small clinically non-relevant differences (for example, LV-E/A). There were no significant differences in laboratory test results between cases with versus without PEM (data not shown).

## Discussion

In this nested population-based case-control study, we found persistence of symptoms and impairments in roughly two-thirds of cases with PCS after more than one year following acute SARS-CoV-2 infection. The comprehensive medical evaluation and comparison of persistent cases with a control group of age- and sex-matched stably convalescent controls demonstrated that many of the persistent cases had objective signs of cognitive deficits and reduced exercise capacity. Apart from observing large and discriminant differences in standardized measures of fatigue, neurocognitive disturbance, sleep quality, perceived stress, depression, anxiety, dysautonomia and quality of life, we detected significant differences between persistent cases and stable controls in MoCA, SDMT and TMT-B tests, in grip strength, VO_2max_, VE/VCO_2_ slope and a few other exercise capacity measures, and this finding was independent of age, sex, BMI and education (as the probably most significant potential confounding variables) and other variables. In contrast, laboratory tests (including inflammatory and coagulation markers) or resting echocardiographic results were not different after adjustment for covariates and were unable to discriminate cases from controls.

### Trajectories

In the majority of participants who had developed PCS after 6 to 12 months following acute SARS-CoV-2 infection, symptoms and complaints persisted for at least another 6 to 12 months. Furthermore, most of the 32% of PCS cases who reported an improvement at follow-up did not fully recover. In a recent Swiss study,[34] the proportion of subjects returning to a normal health status between 6 and 24 months after acute infection was roughly 25%, while the rate of improvement of symptoms associated with PCS was 37%. In another Swiss study[35], the proportion of PCS patients with improvement between months 7 and 15 after acute infection was 48%. In both studies as well as in other work,[36–38] there was a tendency of disease chronification beyond 6 to 12 months after acute infection, and our current findings support these observations. We saw some differential evolution of the predominant symptom clusters between phase 1 and phase 2. Fatigue, chest symptoms, and smell/taste disorders showed a net decrease over time. In contrast, improvement of the cognition and the depression/anxiety/insomnia clusters was similar to aggravation, resulting in only minor changes in the net prevalence. Others have also observed a tendency for more persistence of neurocognitive disturbances rather than other symptom clusters.[16,39–45] Stratified longitudinal analyses with objective measures are needed to better evaluate chronicity and prognosis of cognition deficits or other organic impairments, and such studies may benefit from advanced methods for defining different recovery clusters and multi-parameter modelling with validation across different cohorts.[7,46–49]

Interestingly, risk factors for improvement of case status in the present study included higher educational status, and this was complemented by the finding of lower educational status as a risk factor for worsening health among controls – besides secondary SARS-CoV infection. In the study reported by Hartung and colleagues,[50] lower education was associated with cognitive non-recovery but not with persisting fatigue. In a large online survey[45], lower educational status was associated with worse symptom scores at all time points post-infection, including <24 months. In our previous phase 1 study, lower educational status was already found to be associated with symptomatic disease at 6 to 12 months post-infection, and a similar association has been reported from two large US-cohorts.[51] We cannot exclude that sampling bias accounts for these observations. The fact that we found cases without recent specialist consultation and without participation in rehabilitation between phases 1 and 2 to be more likely to improve, most likely reflects a less severe acute and post-acute illness with a better prognosis (i.e. reverse causation).

An important finding was that post-acute vaccination against SARS-CoV-2 did not appear to be associated with PCS improvement. Several studies have shown a decreased PCS prevalence after vaccination, but it was often unclear[11] whether one or more of the vaccine shots were in fact administered after illness onset. Also, many studies were retrospective and did not adjust for confounders. In the study reported by Tran and colleagues,[52] in which vaccine recipients with PCS were propensity score matched to non-vaccinated individuals with PCS and observed for four months, there were positive associations of (a first) vaccination with symptom severity and remission of PCS. In our study, the proportion of post-infection vaccine recipients was large. Almost all participants had already received their first vaccine shot before phase 1 (without measurable effects on symptom prevalence and severity), and many had received their second or booster doses between phase 1 and phase 2. As almost all had been vaccinated, it is difficult to ascertain a relationship between vaccination and recovery from PCS.

### Symptoms and signs

Symptom ratings and questionnaire data consistently showed that fatigue and cognitive disturbance were the most prevalent health problems (>60% for each cluster) among persistent cases, a finding confirming the results of other studies[41] with a similar follow-up time. Of note were the large overlap between self-reported fatigue, cognition problems and chest symptoms and the correlation of various symptom ratings with health-related quality of life scores. Extreme fatigue and symptoms compatible with ME/CFS affected approximately one-tenth of persistent PCS cases, while PEM lasting >14 h was reported by 36% of persistent cases and was associated with worse scores in all questionnaires, including those on fatigue, sleep, perceived stress, dysautonomia and quality of life, but also in cognitive and cardiopulmonary exercise tests. This underscores the usefulness of including the history and duration of PEM when exploring patients with possible PCS.[53,54] Using the full set of DePaul questionnaire items, estimates for PEM might have been higher. In a Swiss cohort, PEM was observed in 48% of PCS patients, but in that study, fewer subjects (6%) fulfilled the criteria for ME/CFS.[55] A prevalence of 45% for PEM was observed in a Dutch cohort[56] of PCS patients.

Cognitive disturbance was the second most frequent symptom cluster, on the basis of both the symptom reports and the FLei questionnaire ratings, with concentration problems being somewhat more prevalent than memory problems. A similar observation independent of the time after acute infection has also been made in a large online survey[45] among subjects with complaints for at least three months after infection. In a large claims data network analysis of neurologic and psychiatric sequelae, Taquet and colleagues,[57] found that risks of cognitive deficits, dementia, psychotic disorders, and epilepsy/seizures remained increased over a 2-year follow-up period after SARS-CoV-2 infection, which was unlike the risks of common psychiatric disorders that rapidly returned to baseline. Other studies[16,39–45] also reported persisting or increasing cognition or concentration problems with generally decreasing rates of other symptoms and physical health over time. A large memory questionnaire study[58] found increased scores indicating worse memory problems up to 3 years after acute infection (when compared to uninfected controls), and a recent elegant study[59] showed reaction time slowing with increasing time after acute infection. Taken together, these findings and the results of the present study indicate that cognition problems might, in fact, tend more to chronicity than other health problems of PCS patients. Reports of lower prevalences (22-32%) of cognitive disturbances in meta-analyses may be due to differences in sample composition (as the majority of the included studies investigated patients hospitalized during initial disease) and follow-up times.

We found sleep disorder, in particular insomnia, being reported as another frequent complaint among cases. Pooled data of previous studies[60] on >15,000 participants revealed a prevalence of 40 to 50% for sleep disorder among PCS cases, which is comparable to our data. The importance of pre-pandemic healthy sleep to prevent PCS has been demonstrated by us and others.[61,62] It will be interesting to explore whether poor sleep quality remains a risk factor for continued non-recovery from PCS.

Symptom reports and rating data on depressive and anxiety symptoms generally fit in the meta-analyses[60,63] on neuropsychiatric manifestations in PCS. The difference among persistent cases in the proportion reporting depression as a symptom versus having a clinically significant PHQ-9 score might reflect perceived psychological strain rather than having developed a manifest depressive syndrome. With regard to anxiety, the opposite effect was observed: persistent cases subjectively perceived themselves as less anxious than suggested by the anxiety questionnaire score, which may be attributable to an overlap between PCS (nervousness, irritability) and GAD-7 questionnaire items.

We note that most of the routine clinical examination results and laboratory measurements did not discriminate between persistent cases and controls, including resting left ventricular systolic and diastolic function and the Tiffeneau test. These findings are essentially in line with the results of many other groups.[46,64–68] Small differences in values after crude analyses were no longer statistically significant after adjustment, in particular for BMI, smoking status and study site. D-dimer levels, for example, were slightly elevated among persistent cases, but the differences were not significant in adjusted analyses, a result similar to those seen in earlier reports.[65,69,70] Because several studies[20,71,72] suggested hypocortisolism as a possible explanation for PCS in at least some patients, we included blood levels of cortisol, ACTH and DHEA-S in our analysis. However, we could not find significant differences between persistent cases and stable controls, suggesting a low likelihood of subacute or chronic adrenal insufficiency as a major contributing factor for PCS symptoms. Other recent studies[23,73,74] also failed to identify differences in cortisol levels between PCS patients and several control groups. Furthermore, we were not able to detect differences between persistent cases and stable controls in complement turnover, a hypothesis recently raised in a number of studies.[75,76] We did, however, screen only for differences in CH50 between PCS cases versus controls, not for individual complement component blood levels.

Serological investigations indicated that the SARS-CoV-2 spike S1 antibody levels in our cohort were essentially driven by vaccination rather than associated with PCS (as reported by Klein and colleagues),[20] and we did not find a significant association between elevated EA-D IgG antibodies (suggesting EBV reactivation) and PCS in an adjusted analysis. Previous data on this issue have been conflicting, with studies reporting[20,77,78] or failing to report[79,80] EBV reactivation markers associated with PCS. It has to be kept in mind that EA-D IgG antibody levels rise early after active viral replication and typically remain positive for only three to six months, while our samples were collected >12 months after acute SARS-CoV-2 infection. However, we also did not observe increased levels of IgG antibodies against EBNA, which has been suggested as a longer-lasting surrogate for EBV reactivation and that have previously been associated with neurocognitive disturbances in PCS patients. [81]

SARS-CoV-2 persistence has been proposed as another mechanism in non-recovery and PCS development. However, in our analysis, we did not observe antigen positivity in nasopharyngeal specimens, PCR positivity in stool samples, or viral antigen in plasma, which argues against persistent virus replication as a driver of PCS. The prevalence of viral persistence in non-invasive biospecimens from PCS cases as measured by a variety of methods has also been low in previous studies[80,82–84] with the exception of two small studies[85,86] that showed spike antigenemia in >60% of PCS patients some of whom were also PCR-positive in plasma samples and a study reporting S1 protein persistence in monocyte populations of PCS patients up to 15 months post-infection [87]. A recent large study[88] demonstrated that throat swab samples in a subgroup of PCS patients with repeated PCR positivity in the early post-acute phase became negative beyond three months after acute infection. Both spike and N protein were detected in plasma samples of 10 out of 100 patients with severe illness for at least three months (exact times not stated) after COVID-19,[80] but there was no apparent link between detectable antigen and symptoms. No viral RNA was detected in stool samples taken >300 days after acute infection, and prolonged shedding was associated with gastrointestinal symptoms but not PCS. In an exploratory study,[89] 4 out of 5 subjects with a variety of symptoms had positive SARS-CoV-2 RNA detected in rectal biopsies obtained between days 158 and 676 after acute infection.[90] So far, very few patients with PCS and symptoms >12 months have been investigated for antigen/protein and/or RNA persistence,[91] and an association between viral persistence and PCS remains an unproven hypothesis.

### Cognitive deficits

Neurocognitive testing showed significant group differences, indicating cognition deficits concerning attention and executive functioning, with problems in divided attention (TMT-B) and lower processing speed (SDMT) in cases with persistent PCS, and this finding appeared to be independent of pre-existing illnesses. One-third of the persistent cases (versus 18.9% among stable controls) showed MoCA values <26, which is slightly higher than observed in previous studies.[59,92] The mean value among persistent cases was 26.2 (25.8 in cases with PEM) compared with 26.9 among stable controls and similar values among worsening controls and improved cases. This small albeit significant difference may at least partly be related to the fact that the MoCA has limited specificity as a test originally designed to sensitively detect mild cognitive impairment among the elderly.

Impaired executive functioning and reduced processing speed, as observed in our persistent cases is in agreement with a report of similar deficits observed in a large registry cohort[15] of COVID-19 patients followed up with multi-domain cognitive assessment, with pronounced cognitive slowing in 270 patients from two PCS cohorts,[15,59] and with attention and executive function deficits in a comprehensive cognitive assessment of PCS[93] patients after mostly mild initial disease. Although the cognitive findings described in the present study may be insufficient as a diagnostic aid to differentiate cases from controls because of the small to medium effect sizes, the data can help to understand the aetiology of cognitive impairments in PCS. Controlling the group differences in cognitive test results for fatigue or depressive symptoms attenuated the association of the case status with the MoCA to some degree but had little effect on the SDMT and TMT-B group differences, indicating that depressive mood and fatigue alone cannot explain the reduced performance in cognitive tests. This is in accordance with previous data.[94] Taken together, the information so far supports the concept of different pathomechanisms with regard to depression and cognitive disorders in PCS.

### Reduced physical exercise capacity

An impaired exercise capacity with reduced handgrip strength (or 6-minute walk test) and reduced VO_2max_ appear to be hallmark signs of PCS. Both measures were significantly different between persistent PCS cases and stable controls in the present study. A reduced VO_2max_ (<85% predicted) was observed in 35% of the persistent cases, which is comparable to the prevalence found[95] recently in other studies. Similar to earlier observations,[95–99] we also found a lower peak heart rate among persistent cases, while RER_max_ and the rate of perceived exertion were similar. Taken together, these findings are compatible with deconditioning as a major contributor to the impaired performance[100] capacity, but muscular dysfunction/myopathy possibly due to mitochondrial lesions, may be an alternative explanation. Ventilatory inefficiency is likely to be another contributing factor. Breathlessness as a moderate to severe symptom was reported by almost 50% of persistent cases, which also had significantly higher VE/VCO_2_ slope values than stable controls. Other investigators have also found such differences in VE/VCO2 slope between cases and controls.[68,101,102] The prevalence among cases of a VE/VCO_2_ slope >30 (increased) or >34 (abnormal) in our study was substantial (35% and 14%, respectively), greater than among stable controls and similar to the proportions reported by Sorensen[95] and colleagues. Even subtle differences in VE/VCO_2_ slope may impact cardiorespiratory symptom severity[97,99] after exercising. Besides hyperventilation, erratic breathing with high variability in tidal volume and breathing frequency was described in quite a number of PCS patients.[103–107] However, there is no universal gold standard for diagnosing dysfunctional breathing, and the present study did not include systematic screening for erratic breathing. Again, dysfunctional breathing would also be compatible with respiratory muscular dysfunction.

In accordance with previous data,[96,101,108] the normal systolic function in the resting echocardiography in persistent cases described in the present study suggests that the reduced performance capacity is not caused by central cardiac limitation. Also, bronchial obstruction does not seem to be a cause for the hyperventilatory response to exercise since Tiffeneau tests were similar across all subgroups and breathing reserve was not exhausted. The (slightly) reduced FVC among cases (95.9% versus 99.1% for controls) is small but noteworthy.

Longitudinal studies[68,109,110] assessing FVC changes over time after SARS-CoV-2 infection produced conflicting results, while several cross-sectional studies[68,96,111,112] have shown reduced lung volume associated with persistent symptoms. In a study with patients hospitalized for acute infection,[113] reduced FVC at four months correlated with increased findings in chest tomographs, reduced lung diffusion capacity, lower SpO_2_, reduced exercise capacity, more fatigue and lower quality of life. The reason for the lower lung volume in our stable cases who had typically not been hospitalized may be respiratory muscle weakness[107,114,115] which remains to be further elucidated. There has been no clear evidence[97,116] for an impairment of lung diffusion capacity among patients with initially mild acute infection. Lung diffusion capacity was not measured in the present study. However, SpO_2_ at cessation of exercise was not different between groups, making such an hypothesis in our study participants unlikely. Finally we cannot exclude that the CPET results were affected by a lower level of physical fitness already existing prior to infection. The persistent impaired exercise capacity shown here might best be explained by multisystem dysfunction with a peripheral limitation, e.g. impaired oxygen extraction due to mitochondrial dysfunction[117–119] and/or a low preceding fitness level[120] rather than a central cardiac or pulmonary limitation. The roles of dysfunctional breathing and chronotropic incompetence need to be further investigated. In addition, it is not clear what the relatively frequent orthostatic complaints (measured via the COMPASS-31 instrument) contribute to reduced exercise capacity and how this correlates with dysfunctional breathing and chronotropic incompetence.

### Strengths and limitations

One of the strengths of the present study is the nested, population-based approach in defined geographic regions with a large number of subjects with PCR-confirmed earlier infection, regardless of the need for medical treatment. We focussed on adults in the classical working age. We avoided an overrepresentation of hospitalized elderly patients who are likely to show more SARS-CoV-2-nonspecific adverse health sequelae due to more severe acute infection, comorbidities and ageing. We used within-participant comparisons considering symptom frequency before acute SARS-CoV-2 infection and considered only new symptoms not present before the acute infection. In addition, we included at least moderate severity of symptoms and considered impaired activities of daily living or work ability in our working definition of PCS. Another strength is the comprehensive clinical diagnostic work-up of the study participants, including both cases and controls, which included medical history and physical examination, laboratory investigations, CPET and a neuropsychiatric characterization and cognitive assessment. The study allowed us to provide comparative analyses with adjustment for important confounders such as BMI, smoking, and educational level and to stratify the population of persistent PCS cases by the presence of PEM (lasting >14 hours) as a probably important as well as pragmatic and simple surrogate for severity.

An important limitation is that we had no information on exercise capacity before acute infection. We did not perform lung diffusion capacity measurements or neuroimaging and more valid measures of dysautonomia that may provide a more comprehensive understanding of the pathophysiology of PCS. Virological analyses were performed only on serum and – for a representative part of the cohort – on stool samples, but did not include the analysis of biopsy material. Furthermore, the time of sample collection >1 year post SARS-CoV-2 infection may have precluded detection of any transient changes induced in the course of acute infection. Recall bias may be particularly relevant in subjects with more severe neurocognitive deficits. Study participation was higher by cases than by controls from phase 1, and subjects with risk factors (e.g. smoking, obesity) were less likely to respond. Another limitation is the lack of opportunities to include PCS cases with difficulties attending the study centres because of disease severity and who would have needed admission or more support by accompanying relatives or nurses during travelling and outpatient assessment with medical tests. This might also have caused an underestimation of the prevalence of both ME/CFS and longer-lasting PEM. In addition, our screening did not include all DePaul questionnaire item scorings, which may yield PEM prevalence estimates among subjects with PCS of up to 50% or even higher.[67,121–125] We note that the selection of cases fulfilling specific PCS criteria and controls with full recovery after COVID-19 and without complaints and moderate/severe symptoms (i.e. extreme phenotype selection) may lead to higher AUCs of the questionnaires when compared to representative populations. Furthermore, the population is not representative of Germany since we derived our study participants from a population of medium-sized university cities in the southwestern part of the country with substantial sociocultural and socioeconomic differences from other regions in the country. Finally, we did not include subjects from phase 1 who had symptoms compatible with PCS but did not meet the working definition criteria.

### Conclusions and implications

We report that two thirds of PCS cases 6-12 months after acute SARS-CoV-2 infection continue to report persistent symptoms interfering with daily living and associated with reduced quality of life and/or work ability another 6-12 months later. The symptoms appear to change slightly but the predominant symptoms, often clustering together, remain fatigue, cognitive disturbance and chest symptoms, including breathlessness, with sleep disorder and anxiety as additional complaints in a substantial proportion of cases. In a thorough medical examination, many persistent PCS cases show findings that significantly differ from controls and are in part abnormal/out of reference; these include impaired executive functioning, reduced cognitive processing speed and reduced physical exercise capacity only in part explained by deconditioning and typically unrelated to central cardiac or pulmonary limitations. Cases reporting PEM lasting longer than 14 h complained about more severe symptoms and showed worse findings in both cognition and exercise capacity testing. Our findings do not support hypotheses of viral persistence, EBV reactivation, adrenal insufficiency or increased complement turnover as pathophysiologically relevant for persistent PCS.

The results call for the inclusion of cognitive and exercise testing in the clinical evaluation and monitoring of patients with suspected PCS. Together with other research findings, they suggest that further studies should be undertaken to assess the role of skeletal muscle metabolism as well as neurometabolic and neuroinflammatory disorders[126,127] and dysautonomia for an advanced understanding of PCS development and prognosis. Observational studies with longer follow-up are urgently needed to evaluate factors for improvement and non-recovery from PCS.

## Contributors

WVK led the study conceptualisation and the development of the research question, supported by AlN, BFB, BM, CS, JMS, PD, SG, and UM. DR, JS, SG, UM and WVK supervised the study. SG, UM, JMS, PD, BFB, AnN, GE, RG, VG, KK, PM, LM, JS, US, MZ and WVK were responsible for or involved in the clinical work and assessments, and BM, SP, UM, HGK and WVK oversaw the laboratory investigations. RSP, AlN, MR, and DR were involved in data acquisition and statistical analysis. AlN coordinated the biobanking procedures. JMS and JS were responsible for the central evaluation of the echocardiography findings. AnN, BFB, BM, CS, HGK, JS, PD, RSP, SG, UM, and WVK contributed to the analyses and interpretation of the data. WVK, AlN, RSP, and DR had full access to and verified the data, take responsibility for data integrity and the accuracy of the data analysis, and for the decision to submit for publication. All authors were involved in drafting or critically revising the manuscript, and all authors approved the final version. The corresponding author attests that all listed authors meet authorship criteria and that no others meeting the criteria have been omitted.

## Funding

This work was funded by the Baden-Württemberg Federal State Ministry of Science and Art (grant number MR/S028188/1).

## Declaration of interests

We declare no competing interests.

## Ethical approval

Ethical approval was obtained from the respective ethical review boards of the study centres in Freiburg (21/1484_1), Heidelberg (S-846/2021), Tübingen (845/2021BO2), and Ulm (337/21).

## Data sharing

Data from EPILOC phase 2 areavailable for research purposes upon reasonable request. The study guarantor (WVK) affirms that the manuscript is an honest, accurate, and transparent account of the study being reported; that no important aspects of the study have been omitted; and that any discrepancies from the study as planned have been explained.

## Data Availability

All data produced in the present study are available upon reasonable request to the authors.

## Acknowledgements

We thank all study participants with their care-givers and key collaborators (in alphabetic order) on this work: Julian Böhm, Stefan Brockmann, Stefanie Bröer, Christof Burgstahler, Katharina Caesar, Bettina Deibert, Xiaohong Du, Nelli Edel, Sabine Gerbersdorf, Jennifer Hermann, Katja Hirth, Achim Jerg, Johannes Kirsten, Manuela Licka, Jennifer Müller, Hasema Persch, Patrick Roling, Stephan Rusch, Michaela Schmid, Patrick Schneeweiß, Katarina Stete, Elisabeth Stoll, Adrian Tassoni, Hanna Tschischka, Shirin Vollrath, Vanessa Walz, Dietrich Walzer. We acknowledge the participating local laboratories and biobanking facilities for their technical support.

## Notes

### Competing Interest Statement

The authors have declared no competing interest.

### Funding Statement

This work was funded by the Baden-Wuerttemberg Federal State Ministry of Science and Art (grant number MR/S028188/1).

### Author Declarations

Ethical approval was obtained from the Ethics Committee of the University of Freiburg, Engelberger Strasse 21, D-79106 Freiburg/Germany (#21/1484_1), the Ethics Committee of the Medical Faculty of Heidelberg University, Alte Glockengiesserei 11/1, D-69115 Heidelberg/Germany (#S-846/2021), the Ethics Committee at the Medical Faculty of the Eberhard-Karls-University and at the University Hospital of Tuebingen, Gartenstrasse 47, D-72074 Tuebingen/Germany (#845/2021BO2), and the Ethic Committee of the University of Ulm, Oberberghof 7, D-89081 Ulm/Germany (#337/21).

