## Supplementary tables & figures for "Persistent symptoms and clinical findings in adults with post-acute sequelae of COVID-19/post-COVID-19 syndrome in the second year after acute infection: population-based, nested case-control study"

### **Supplementary tables and figures** (results including sensitivity analyses)

**Supplementary figure S11.** Sensitivity analysis 1, excluding participants with health conditions already present before index infection (cardiovascular diseases, respiratory diseases, mental disorders, neurologic or sensory disorders, cancer, metabolic diseases, n=599) and cases with a possible alternative medical explanation of persisting symptoms (n=41). Shown are means (geometric mean for COMPASS-31 and TMT-B) of self-reported health outcomes and neurocognitive tests (with 95%-CI) by case-control status at clinical examination in phase 2 ....... 18

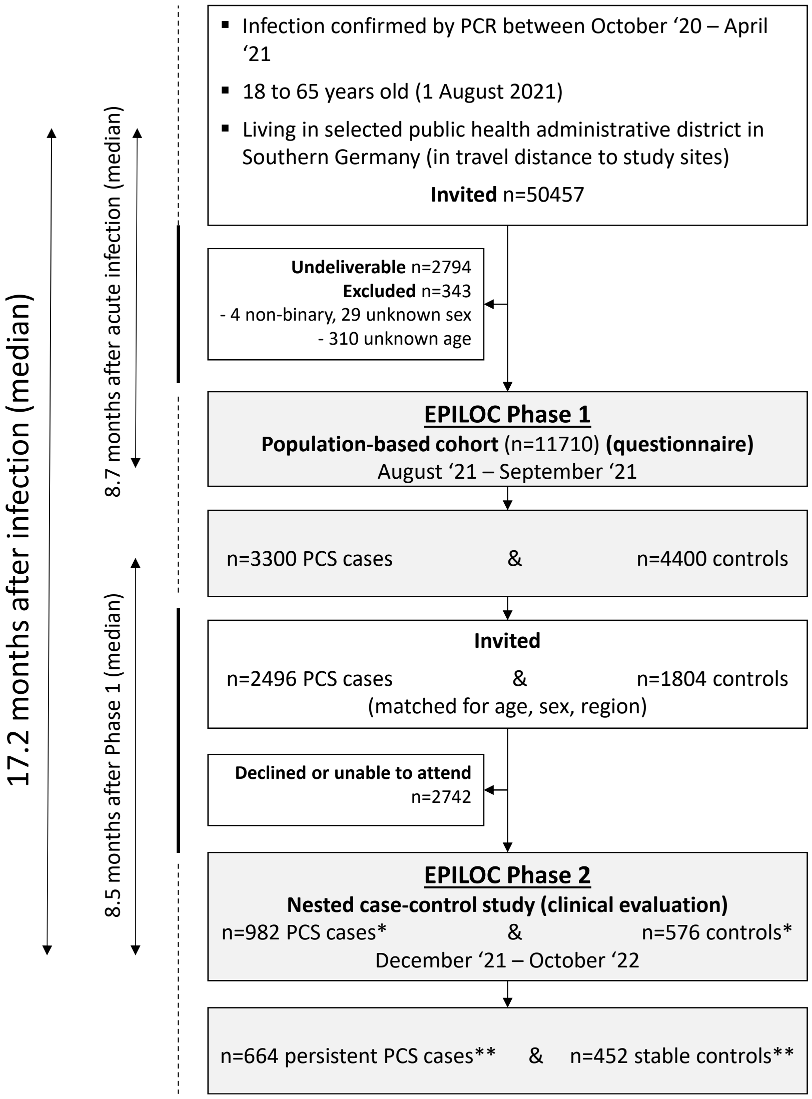

*before (at phase 1) and ** after clinical examination (in phase 2)

**Supplementary figure S1.** Flowchart covering EPILOC study phases 1 and 2.

**Supplementary table S1.** Characteristics of case and controls from phase 1 who participated in the phase 2 clinical examination.

| | Phase 1 cases | | Phase 1 controls | |
|---|---|---|---|---|
| | N | Mean or frequency | N | Mean or frequency |
| ***Phase 1 characteristics*** | | | | |
| Female, N (%) | | 633 (64.5) | | 379 (65.8) |
| Age (years), mean (sd) | 982 | 48.1 (12.3) | 576 | 48.5 (12.3) |
| University entrance qualification, N (%) | 982 | 420 (42.8) | 576 | 338 (58.7) |
| Married/living together | 977 | 847 (86.7) | 574 | 509 (88.7) |
| Never Smoker, N (%) | 979 | 629 (63.7) | 576 | 420 (73.0) |
| Obese (≥30 kg/m²), N (%) | | 225 (23.0) | | 58 (10.1) |
| Full-time employment, N (%) | 981 | 500 (51.0) | 575 | 289 (50.3) |
| Treatment of acute SARS-CoV-2 infection, N (%) | | | | |
| No medical care/treatment | 968 | 541 (55.9) | 573 | 516 (90.1) |
| Outpatient care | | 350 (36.2) | | 49 (8.6) |
| Inpatient care (without ICU) | | 62 (6.4) | | 6 (1.1) |
| Intensive care | | 15 (1.6) | | 2 (0.4) |
| Time from positive PCR test to phase 1 (months), mean (sd) | 980 | 8.4 (1.6) | 571 | 8.6 (1.6) |
| Received first SARS-CoV-2 vaccine, N (%) prior to phase 1 | 981 | 833 (84.9) | 575 | 501 (87.1) |
| Preexisting condition/comorbidities, N (%) | | | | |
| Musculoskeletal disorders (including rheumatism) | 969 | 429 (44.3) | 575 | 170 (29.6) |
| Cardiovascular disorders (including hypertension) | 974 | 223 (22.9) | 576 | 73 (12.7) |
| Neurological or sensory disorders | 978 | 225 (23.0) | 576 | 90 (15.6) |
| Metabolic disorders | 981 | 230 (23.5) | 575 | 88 (15.3) |
| Mental disorders | 976 | 185 (19.0) | 575 | 39 (6.8) |
| Respiratory diseases | 975 | 163 (16.7) | 576 | 48 (8.3) |
| Dermatological diseases | 977 | 130 (13.3) | 574 | 65 (11.3) |
| Cancer | 957 | 39 (4.1) | 560 | 26 (4.6) |
| ***History from phase 1 to phase 2*** | | | | |
| Time from phase 1 to phase 2 (months), mean (sd) | 982 | 9.1 (2.6) | 576 | 8.4 (2.7) |
| Secondary SARS-CoV-2 infection since phase 1, N (%) | 982 | 230 (23.4) | 576 | 134 (23.3) |
| Total number of vaccine shots received, N (%) | | | | |
| 0 | 981 | 33 (3.4) | 577 | 33 (5.7) |
| 1 | | 106 (10.8) | | 39 (6.8) |
| 2 | | 752 (76.7) | | 446 (77.3) |
| 3 | | 90 (9.2) | | 59 (10.2) |
| Participation in post-COVID-rehabilitation program, N (%) | 941 | 90 (9.6) | 565 | 0 (0.0) |
| Physician consultations within six months prior to phase 2, N (%) | | | | |
| None | 982 | 265 (27.0) | 576 | 335 (58.2) |
| General practitioner | | 341 (34.7) | | 193 (33.5) |
| Any specialist physician | | 469 (47.8) | | 56 (9.7) |
| Cardiology | | 178 (18.1) | | 15 (2.6) |
| Respiratory Medicine | | 214 (21.8) | | 18 (3.1) |
| Neurology | | 88 (9.0) | | 1 (0.2) |
| Radiology | | 85 (8.7) | | 8 (1.4) |
| Rheumatology | | 15 (1.5) | | 2 (0.4) |
| Otorhinolaryngology | | 44 (4.5) | | 2 (0.4) |

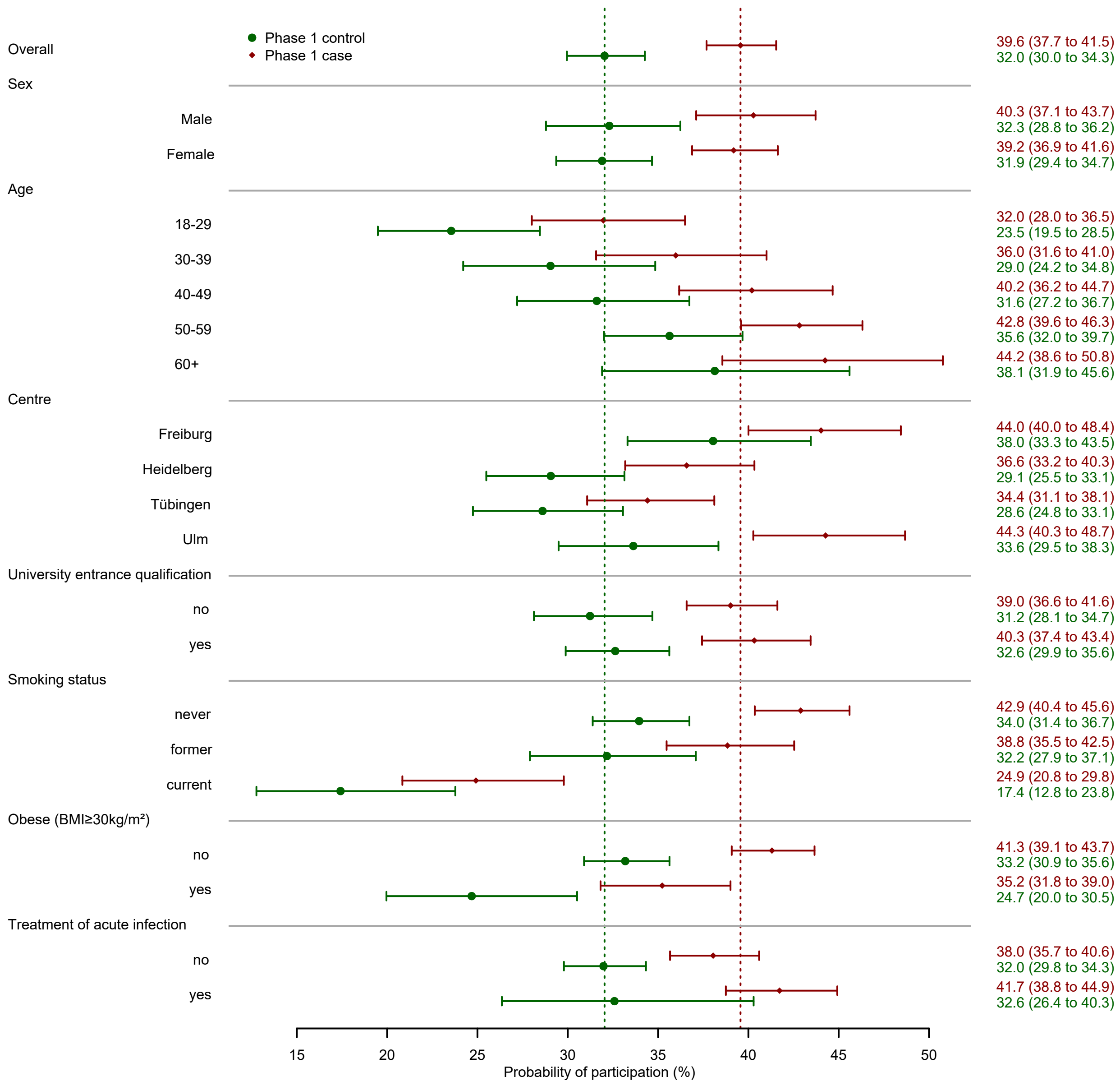

**Supplementary figure S2.** Probability of participation (95% CI) in invited subjects by selected phase 1 characteristics.

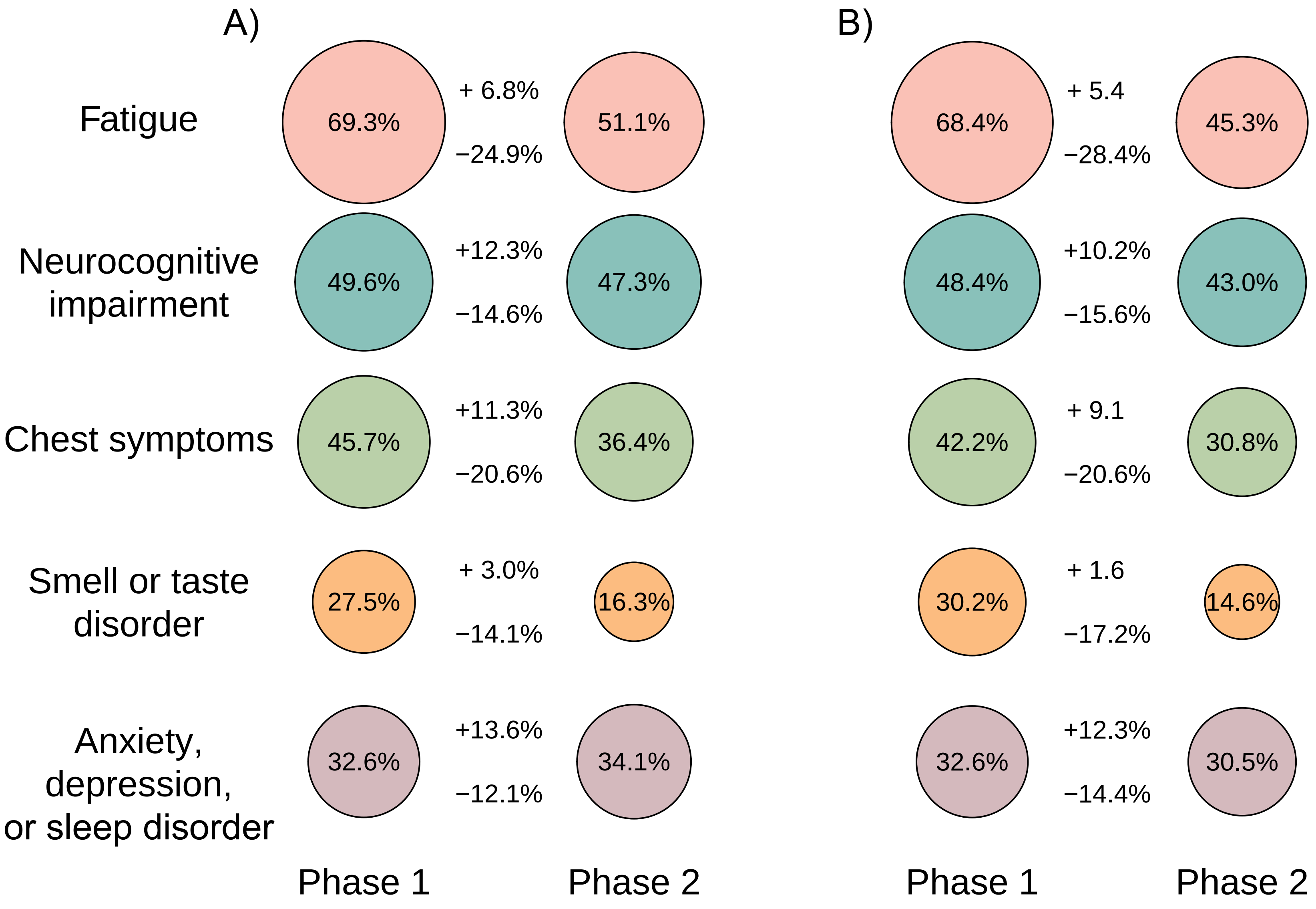

**Supplementary figure S3.** Changes in the prevalence of the five main symptom clusters (based on self-reported new symptoms of moderate to strong severity after acute infection) in phase 1 cases participating in phase 2. A) all participants. B) excluding participants with health conditions already present before index infection (cardiovascular diseases, respiratory diseases, mental disorders, neurologic or sensory disorders, cancer, metabolic diseases, n=599) and cases with an alternative explanation of persisting symptoms (n=41).

**Supplementary table S2**. Mutually adjusted predictors of case-control status change between phase 1 and phase 2.

| **Phase 1 characteristics/variable** | **OR (95%-CI)** | |
|---|---|---|
| **Cases *(N=953)*** | **Persistent Cases (N=613)** | **Cases improved (N=297)** |
| Female sex | 1.00 | 1.15 (0.82 to 1.60) |
| Age (per 10 years) | 1.00 | 0.92 (0.80 to 1.04) |
| University entrance qualification | 1.00 | **1.38 (1.01 to 1.89)** |
| Not married/living together | 1.00 | 0.90 (0.64 to 1.28) |
| Full-time employment | 1.00 | **1.93 (1.39 to 2.67)** |
| Medical care/treatment of acute infection | 1.00 | **0.68 (0.51 to 0.92)** |
| Obesity (BMI ≥ 30kg/m²) | 1.00 | 0.73 (0.51 to 1.06) |
| Time from phase 1 to phase2 (per month) | 1.00 | 1.01 (0.96 to 1.07) |
| Secondary SARS-CoV-2 infection since Ph1 | 1.00 | 1.01 (0.71 to 1.44) |
| Two or more vaccine shots received | 1.00 | 0.87 (0.58 to 1.32) |
| Participation in a post-COVID-19-rehabilitation program | 1.00 | **0.41 (0.21 to 0.79)** |
| Any specialist consultation in the last 6 months | 1.00 | **0.60 (0.45 to 0.81)** |
| | **OR (95%-CI)** | |
| **Controls *(N=573)*** | **Stable Controls (N=445)** | **Controls worsened (N=123)** |
| Female sex | 1.00 | 1.01 (0.62 to 1.62) |
| Age (per 10 years) | 1.00 | 0.96 (0.79 to 1.17) |
| University entrance qualification | 1.00 | **0.53 (0.34 to 0.83)** |
| Not married/living together | 1.00 | 1.26 (0.73 to 2.15) |
| Full-time employment | 1.00 | 1.13 (0.72 to 1.79) |
| Medical care/treatment of acute infection | 1.00 | 1.31 (0.69 to 2.51) |
| Obesity | 1.00 | 1.62 (0.85 to 3.07) |
| Time from phase 1 to phase2 (per month) | 1.00 | 0.94 (0.86 to 1.02) |
| Secondary SARS-CoV-2 infection since Ph1 | 1.00 | **1.85 (1.12 to 3.06)** |
| Two or more vaccine shots received | 1.00 | 1.19 (0.62 to 2.29) |

**Supplementary table S3.** Current medication by case-control status as reported at clinical examination in phase 2.

| | Persistent cases | | Cases improved | | Controls worsened | | Stable controls | |
|---|---|---|---|---|---|---|---|---|
| | N | Mean or frequency | N | Mean or frequency | N | Mean or frequency | N | Mean or frequency |
| Current medication - number of different drugs, N (%) | | | | | | | | |
| None | 664 | 191 (28.8) | 318 | 128 (40.3) | 124 | 53 (42.7) | 452 | 239 (52.9) |
| More than two | | 229 (34.5) | | 60 (18.9) | | 18 (14.5) | | 55 (12.2) |
| Current medication - ATC-Groups, N (%) | | | | | | | | |
| Agents acting on the renin-angiotensin system (C09) | 664 | 134 (20.2) | 318 | 58 (18.2) | 124 | 22 (17.7) | 452 | 51 (11.3) |
| Thyroid therapy (H03) | | 136 (20.5) | | 50 (15.7) | | 14 (11.3) | | 63 (13.9) |
| Vitamins (A11) | | 106 (16.0) | | 25 (7.9) | | 12 (9.7) | | 31 (6.9) |
| Vitamin D (A11CC) | | 91 (13.7) | | 18 (5.7) | | 9 (7.3) | | 28 (6.2) |
| Drugs for obstructive airway diseases (R03) | | 75 (11.3) | | 29 (9.1) | | 8 (6.5) | | 15 (3.3) |
| Lipid modifying agents (C10) | | 71 (10.7) | | 16 (5.0) | | 5 (4.0) | | 31 (6.9) |
| Psychoanaleptics (N06) | | 78 (11.8) | | 15 (4.7) | | 7 (5.7) | | 9 (2.0) |
| Beta blocking agents (C07) | | 71 (10.7) | | 13 (4.1) | | 3 (2.4) | | 14 (3.1) |
| Analgesics (N02) | | 61 (9.2) | | 14 (4.4) | | 8 (6.5) | | 15 (3.3) |
| Drugs for peptic ulcer and gastro-oesophageal reflux disease (A02B) | | 59 (8.9) | | 18 (5.7) | | 4 (3.2) | | 8 (1.8) |
| Calcium channel blockers (C08) | | 41 (6.2) | | 10 (3.1) | | 3 (2.4) | | 19 (4.2) |
| Diuretics (C03) | | 37 (5.6) | | 10 (3.1) | | 2 (1.6) | | 12 (2.7) |
| Antithrombotic agents (B01) | | 38 (5.7) | | 6 (1.9) | | 1 (0.8) | | 12 (2.7) |
| Drugs used in diabetes (A10) | | 31 (4.7) | | 8 (2.5) | | 3 (2.4) | | 5 (1.1) |
| Antihistamines for systemic use (R06) | | 22 (3.3) | | 9 (2.8) | | 5 (4.0) | | 8 (1.8) |
| Antiinflammatory and antirheumatic products (M01) | | 20 (3.0) | | 5 (1.6) | | 2 (1.6) | | 9 (2.0) |

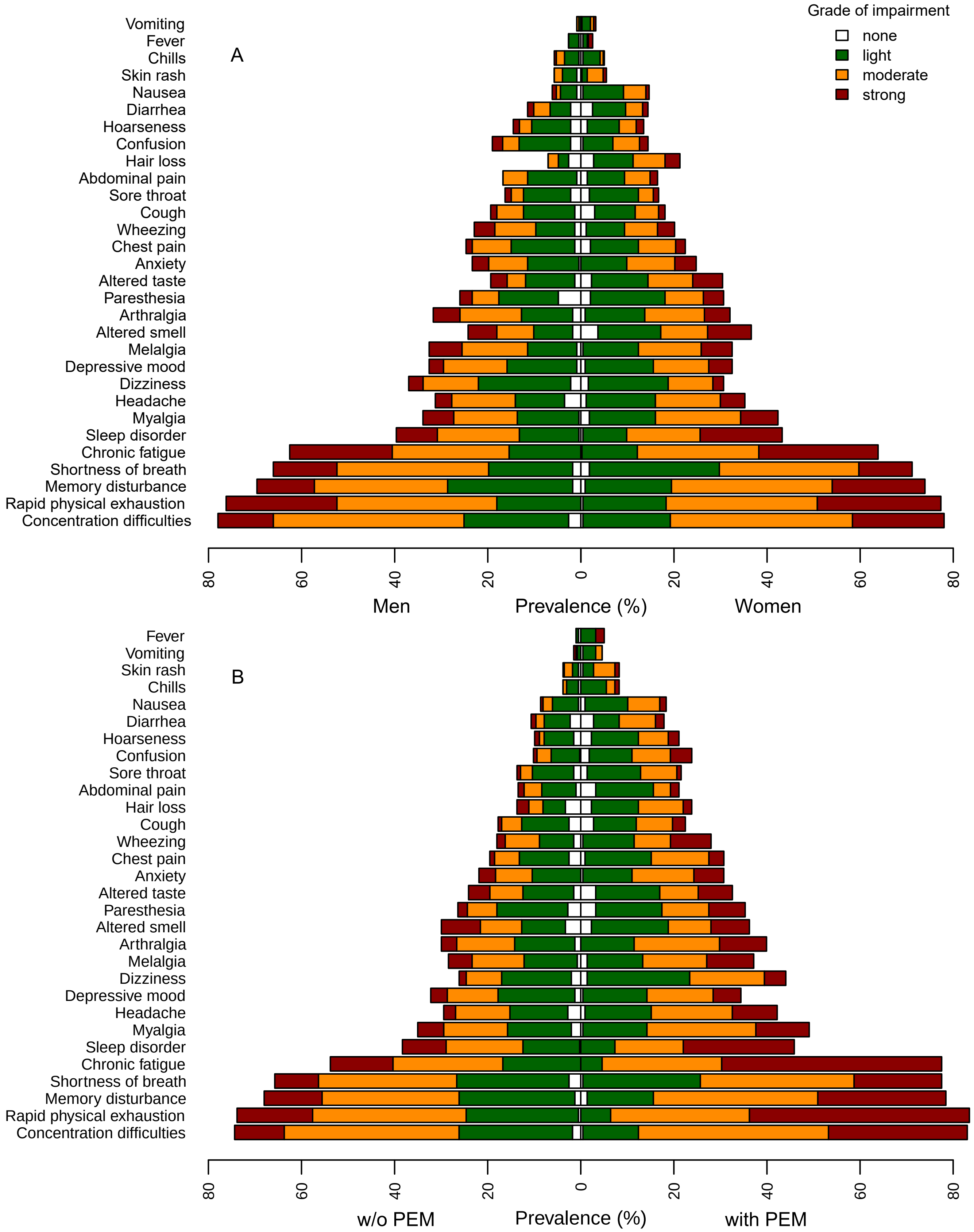

**Supplementary figure S4.** Individual symptoms of different grades among (A) male and female persistent cases, (B) among persistent cases with or without post-exertional malaise (PEM, lasting >14 hours).

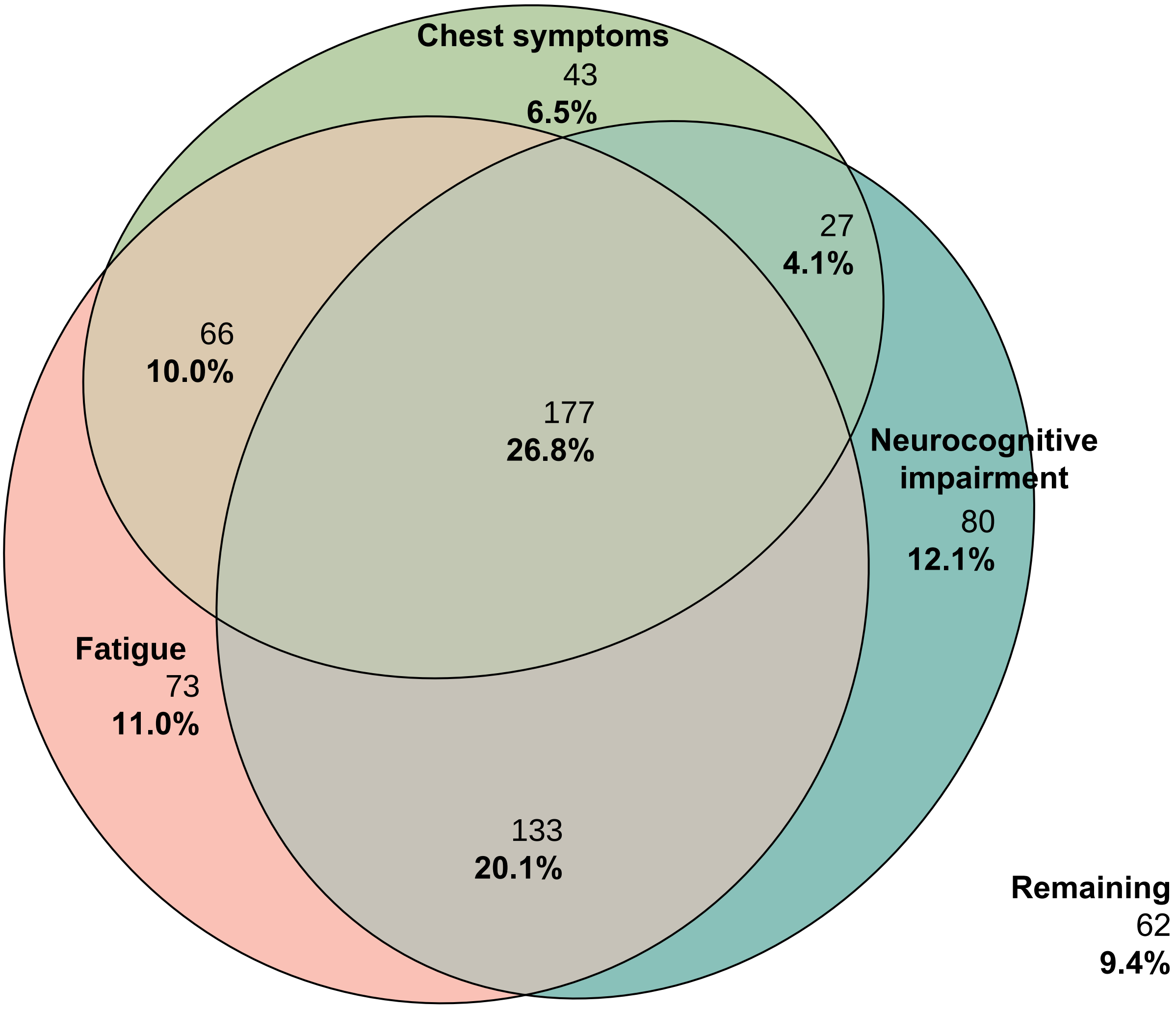

**Supplementary figure S5:** Euler graphs showing the overlap of the three main symptom clusters from phase 2 based on symptoms of grade moderate to strong in persistent cases only.

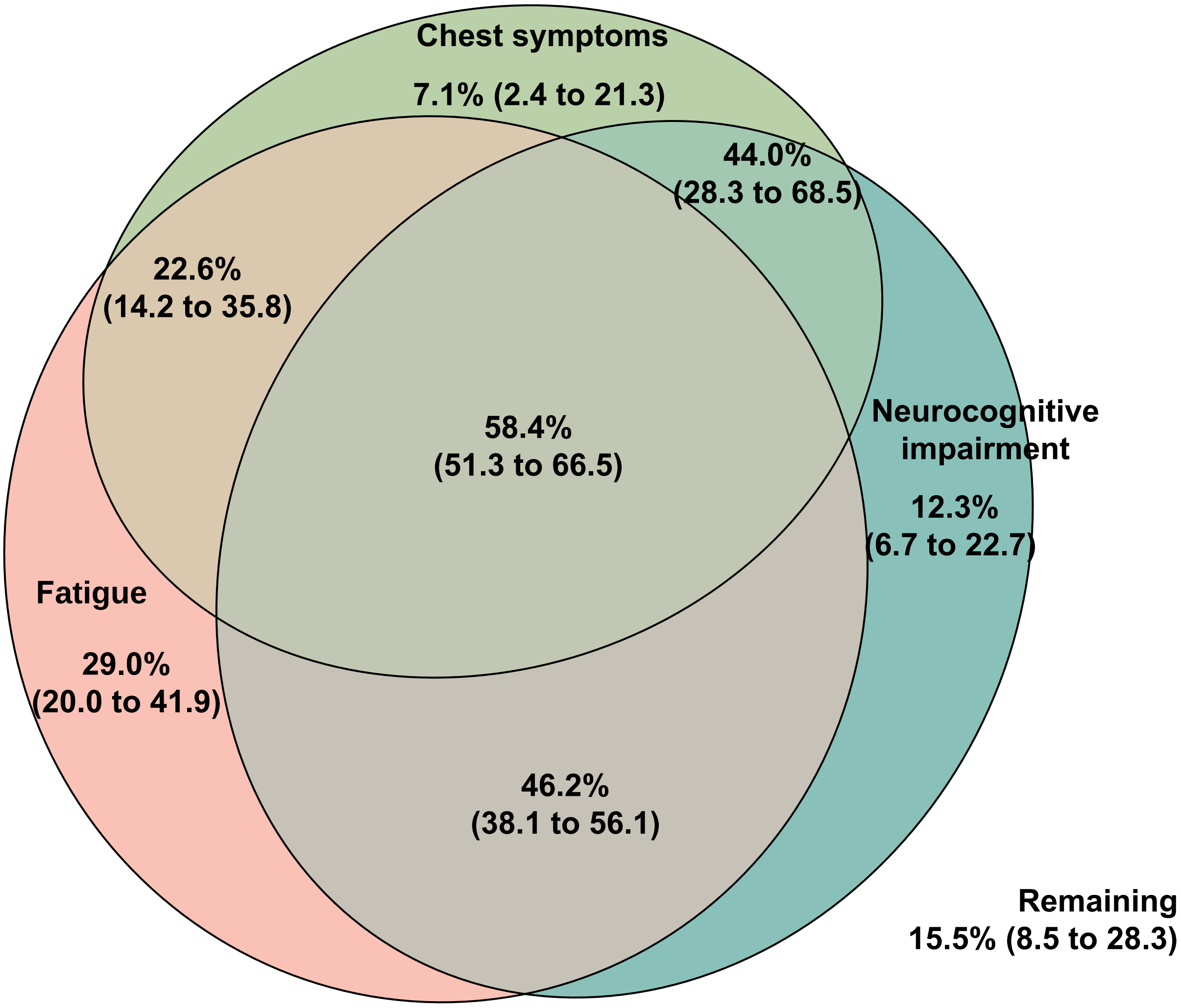

**Supplementary figure S6:** Euler graphs showing the descriptive prevalence (with 95%CI) of post exertional malaise (PEM, lasting >14 hours) in the various overlaps of the three main symptom clusters from phase 2 based on symptoms of grade moderate to strong in persistent cases only.

**Supplementary table S4.** Additional results of resting heart ultrasound examination and CPET analyses by case-control status as reported at clinical examination in phase 2.

| | Persistent cases | | | Cases improved | | | Controls worsened | | | Stable controls | | |
|---|---|---|---|---|---|---|---|---|---|---|---|---|
| | N | Frequency | $OR_{adj}$[1] | N | Frequency | $OR_{adj}$[1] | N | Frequency | $OR_{adj}$[1] | N | Frequency | $OR_{adj}$[1] |
| Diastolic dysfunction grade 1/2 | 608 | 188 (30.9) | 0.93 (0.67 to 1.30) | 294 | 69 (23.5) | 0.87 (0.59 to 1.29) | 115 | 34 (29.6) | 1.11 (0.67 to 1.84) | 424 | 93 (21.9) | 1.00 (ref.) |
| FEV1/FVC <70% of predicted | 551 | 57 (10.3) | 1.08 (0.68 to 1.71) | 273 | 23 (8.4) | 0.78 (0.68 to 1.71) | 105 | 11 (10.5) | 1.01 (0.49 to 2.10) | 384 | 37 (9.6) | 1.00 (ref.) |
| $VE/VCO_2$ slope | | | | | | | | | | | | |
| >30 | | 194 (34.9) | 2.28 (1.64 to 3.17) | | 84 (30.1) | 1.78 (1.22 to 2.58) | | 24 (22.9) | 1.25 (0.73 to 2.15) | | 73 (18.5) | 1.00 (ref.) |
| >34 | | 75 (13.5) | 2.98 (1.67 to 5.32) | | 26 (9.3) | 2.14 (1.11 to 4.12) | | 6 (5.7) | 1.19 (0.44 to 3.18) | | 16 (4.1) | 1.00 (ref.) |
| $VO_{2max}$ (ml/min/kg) | 556 | | | 279 | | | 105 | | | 394 | | |
| <35 in men or <27 in women | | 405 (72.8) | 2.43 (1.74 to 3.39) | | 168 (60.2) | 1.92 (1.32 to 2.81) | | 63 (60.0) | 1.80 (1.07 to 3.06) | | 165 (41.9) | 1.00 (ref.) |
| <20 in men or <17 in women | | 69 (12.4) | 9.44 (2.86 to 31.1) | | 12 (4.3) | 4.34 (1.17 to 16.1) | | 2 (1.9) | 2.02 (0.32 to 12.8) | | 3 (0.8) | 1.00 (ref.) |
| $VO_{2max}$ <85% of predicted | | 196 (35.3) | 5.16 (3.29 to 8.07) | | 63 (22.6) | 3.12 (1.88 to 5.20) | | 13 (12.4) | 1.41 (0.67 to 2.98) | | 33 (8.4) | 1.00 (ref.) |

[1] Adjusted for sex-age class combinations, study centre, university entrance qualification, smoking status and use of beta blocking agents.

**Supplementary table S5.** Neurocognitive tests by CPET results in persistent cases reported at clinical examination in phase 2.

0

| | MoCA | | | SDMT | | | TMT-B | | |
|---|---|---|---|---|---|---|---|---|---|
| | N | ≤25, N (%) | OR[3] | N | < 36[1], N (%) | OR[3] | N | > 109 sec[2], N (%) | OR[3] |
| FEV1/FVC | | | | | | | | | |
| ≥ 70% | 493 | 164 (33.3) | 1.00 (ref.) | 493 | 63 (12.8) | 1.00 (ref.) | 493 | 71 (14.4) | 1.00 (ref.) |
| < 70% | 56 | 20 (35.7) | 0.79 (0.41 to 1.53) | 55 | 7 (12.7) | 0.72 (0.27 to 1.90) | 56 | 7 (12.5) | 0.65 (0.26 to 1.66) |
| VE/$VCO_2$ slope | | | | | | | | | |
| ≤ 30 | 360 | 104 (28.9) | 1.00 (ref.) | 359 | 43 (12.0) | 1.00 (ref.) | 360 | 48 (13.3) | 1.00 (ref.) |
| > 30 | 194 | 81 (41.8) | 1.53 (1.03 to 2.28) | 194 | 27 (13.9) | 1.08 (0.62 to 1.90) | 194 | 31 (16.0) | 1.02 (0.60 to 1.74) |
| > 34 | 75 | 40 (53.3) | 2.06 (1.19 to 3.57) | 75 | 14 (18.7) | 1.80 (0.86 to 3.74) | 75 | 12 (16.0) | 0.88 (0.42 to 1.87) |
| $VO_2$ max, N (%) | | | | | | | | | |
| ≥ 85% of predicted | 358 | 120 (33.5) | 1.00 (ref.) | 358 | 43 (12.0) | 1.00 (ref.) | 358 | 54 (15.1) | 1.00 (ref.) |
| < 85% of predicted | 196 | 65 (33.2) | 1.03 (0.63 to 1.69) | 195 | 27 (13.9) | 2.36 (1.14 to 4.86) | 196 | 25 (12.8) | 1.13 (0.58 to 2.22) |

[1] 15%-percentile of stable controls

[2] 85%-percentile of stable controls

[3] adjusted for sex-age class combinations, study centre, university entrance qualification, BMI, smoking status, and use of beta blocking agents

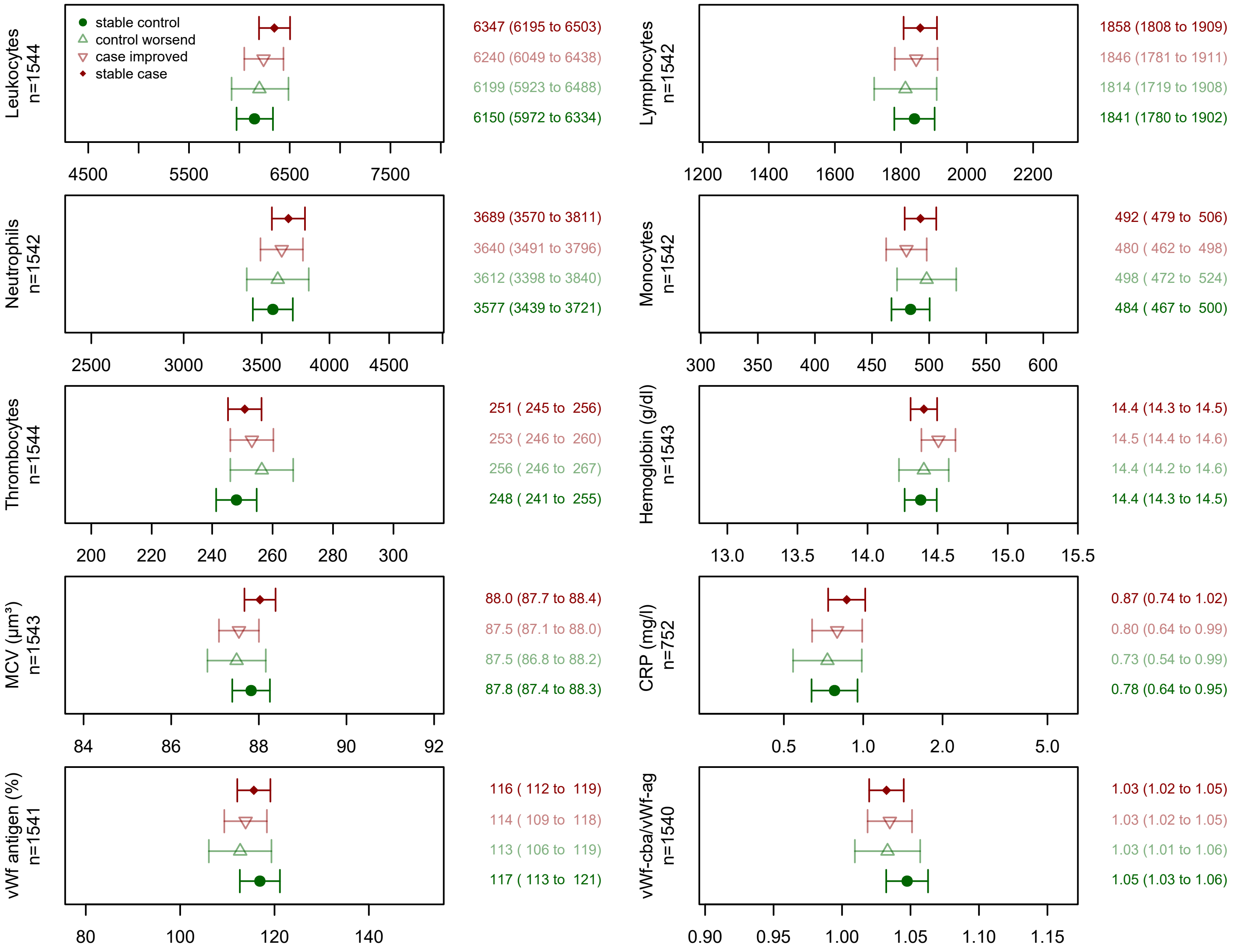

**Supplementary figure S7.** Means (geometric mean for CRP) of blood cell counts (with 95%-CI) by stable case-control status at clinical examination in phase 2. Adjusted for sex-age class combinations, study centre, university entrance qualification. For comparability the x-axis is scaled from mean -1 SD to mean +1 SD for all panels.

**Supplementary table S6.** Case-control status by D-dimer levels (normal vs elevated).

| | D-dimer | | $OR_{adjusted}$* |
|---|---|---|---|
| | ≤0.25 mg/L FEU | >0.25 mg/L FEU | |
| | N (%) | N (%) | |
| Stable controls | 279 (62.1) | 170 (37.9) | 1.00 (ref.) |
| Controls worsened | 72 (58.1) | 52 (41.9) | 1.05 (0.69 to 1.61) |
| Cases improved | 178 (56.3) | 138 (43.7) | 1.10 (0.81 to 1.50) |
| Persistent cases | 342 (52.2) | 313 (47.8) | 1.21 (0.93 to 1.57) |

*Adjusted for sex-age class combinations, study centre, university entrance qualification, BMI and smoking status

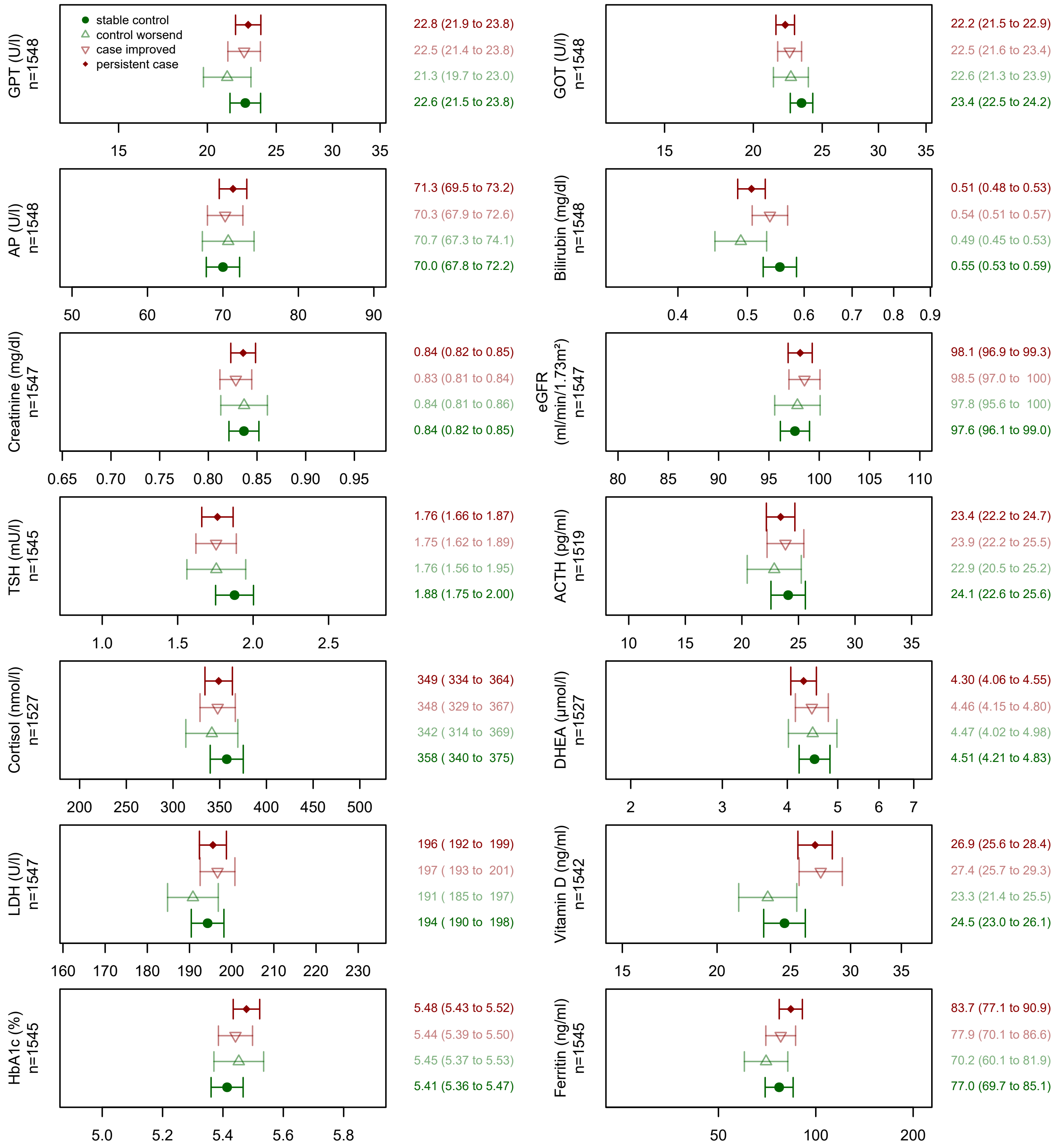

**Supplementary figure S8.** Mean (geometric mean for GPT, GOT, bilirubin, DHEA-S, vitamin D, and ferritin) of selected laboratory measurements (with 95%-CI) by stable case-control status at clinical examination in phase 2. Adjusted for sex-age class combinations, study centre, university entrance qualification, BMI and smoking status. ACTH, cortisol and DHEA-S were additionally adjusted for time of sampling; and vitamin D for intake of vitamin D supplements. For comparability the x-axis is scaled from mean -1 SD to mean +1 SD for all panels.

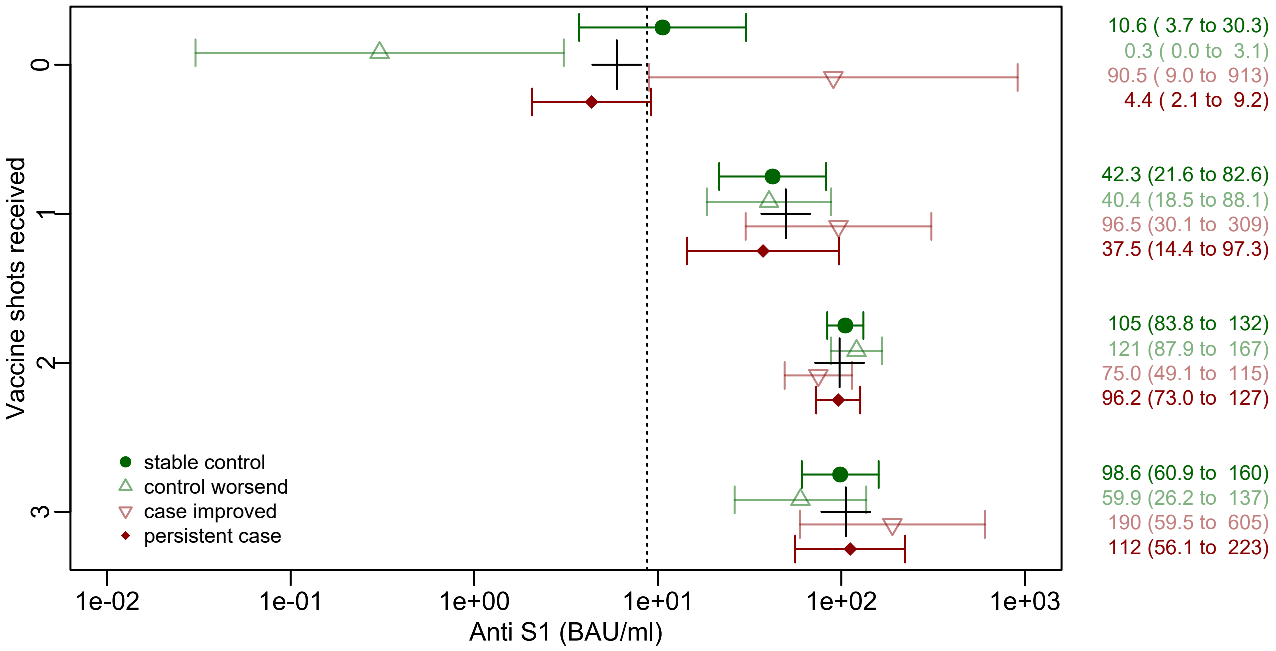

**Supplementary figure S9.** Geometric mean of anti S1 titer (BAU/ml) by number of received vaccine shots and case-control status at clinical examination in phase 1, adjusted for sex-age class combinations, study centre, and university entrance qualification. Quantitative data were determined by dilution of samples for one centre only (N=398). Values left of the dotted line (below 8.75 BAU/ml) are considered negative; crosses represent the geometric mean per number of received vaccine shots independent of case-control status.

**Supplementary table S7.** Phase 2 case-control status by EBV and CMV antibody pattern.

| | EBV antibodies | | | | CMV antibodies | | | |
|---|---|---|---|---|---|---|---|---|
| | N (%) | | | $OR_{Adj}$ [4] | N (%) | | | $OR_{Adj}$ [4] |
| | Negative[1] | Previously infected[2] | Re-activated[3] | Reactivated vs. previously infected | Negative <22 RE/ml | Positive 23-199 RE/ml | Positive ≥200 RE/ml | Positive 23-199 vs. ≥200 RE/ml |
| Stable controls | 22 (4.9) | 372 (83.2) | 53 (11.9) | 1.00 (ref.) | 271 (60.6) | 146 (32.7) | 30 (6.7) | 1.00 (ref.) |
| Controls worsened | 5 (4.0) | 106 (86.2) | 12 (9.8) | 0.77 (0.40 to 1.51) | 78 (63.4) | 35 (28.5) | 10 (8.1) | 1.31 (0.57 to 2.99) |
| Cases improved | 15 (4.9) | 258 (83.5) | 36 (11.7) | 1.00 (0.63 to 1.58) | 189 (61.0) | 99 (31.9) | 22 (7.1) | 1.11 (0.60 to 2.05) |
| Persistent cases | 24 (3.7) | 538 (82.4) | 93 (14.2) | 1.20 (0.83 to 1.74) | 380 (57.9) | 218 (33.2) | 58 (8.8) | 1.27 (0.77 to 1.10) |

[1] Negative: VCA <22 RE/ml and EBNA <22 RE/ml and EA-D <22 RE/ml

[2] Previously infected: (VCA ≥22 RE/ml or EBNA ≥22 RE/ml) and EA-D <22 RE/ml

[3] Reactivated: (VCA ≥22 RE/ml or EBNA ≥22 RE/ml) and EA-D ≥22 RE/ml

[4] Adjusted for sex-age class combinations, study centre, and university entrance qualification

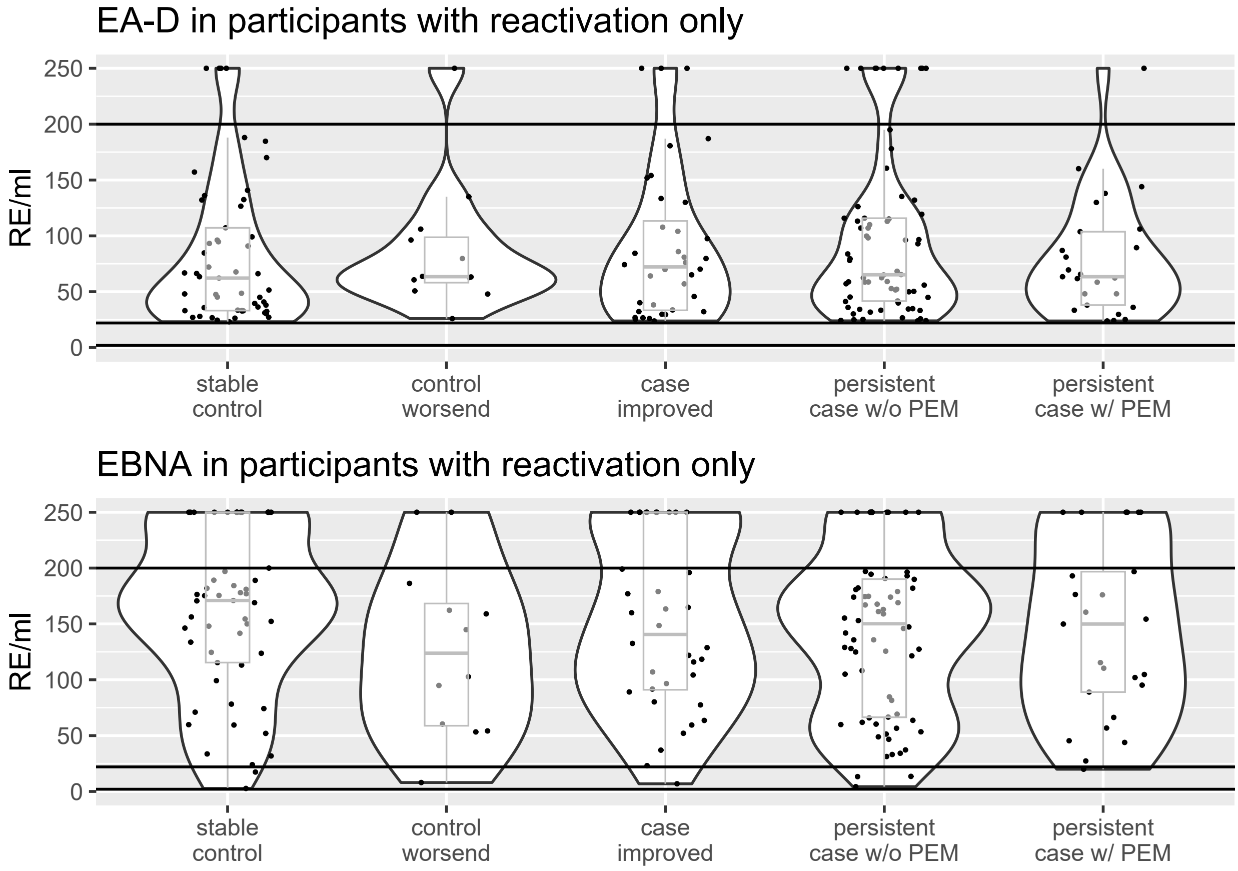

**Supplementary figure S10**. EA-D and EBNA IgG antibody levels in participants with evidence for EBV reactivation.

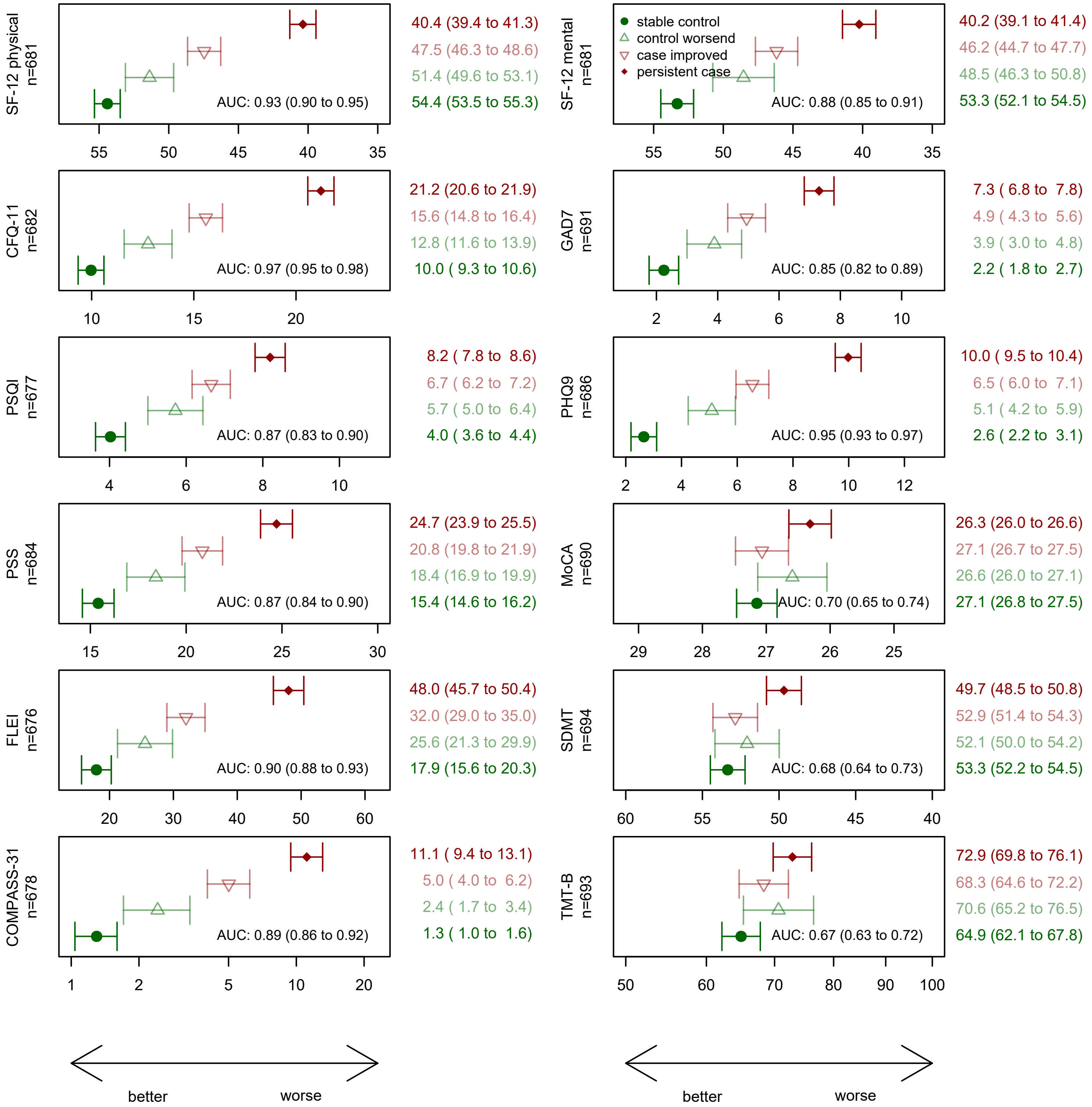

**Supplementary figure S11.** Sensitivity analysis 1, excluding participants with health conditions already present before index infection (cardiovascular diseases, respiratory diseases, mental disorders, neurologic or sensory disorders, cancer, metabolic diseases, n=599) and cases with a possible alternative medical explanation of persisting symptoms (n=41). Shown are means (geometric mean for COMPASS-31 and TMT-B) of self-reported health outcomes and neurocognitive tests (with 95%-CI) by case-control status at clinical examination in phase 2, adjusted for sex-age class combinations, study centre, and university entrance qualification. The reported area under the curve (AUC) for persistent cases vs. stable controls by the respective instrument is adjusted for sex-age class combinations and university entrance qualification. For comparability the x-axis is scaled from mean -1 SD to mean +1 SD for all panels. MoCA: Montreal cognitive assessment scale (points); SDMT: Symbol Digit Modalities Test (number of correct symbols); TMT-B: Trail making test B (time in seconds).

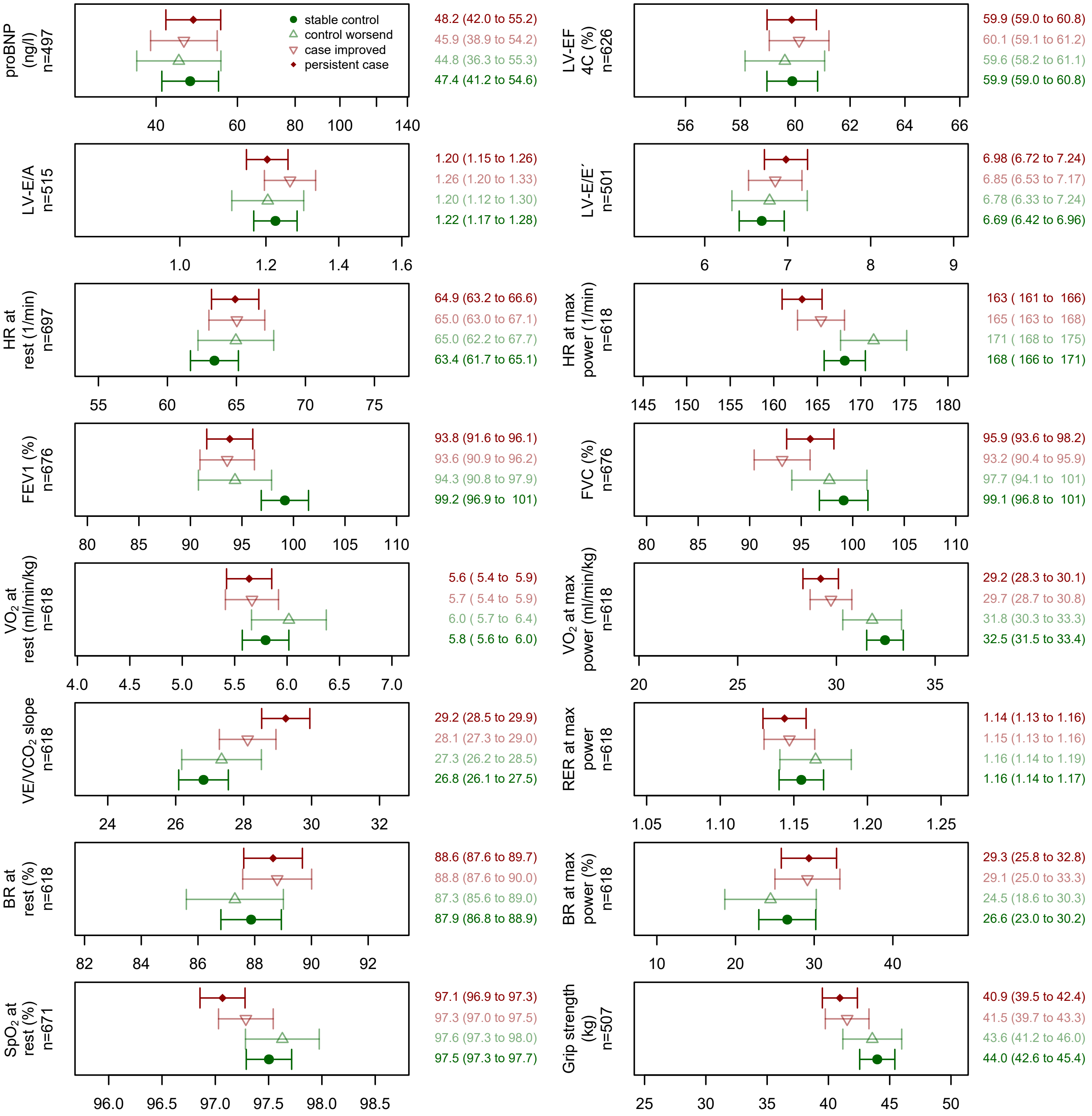

**Supplementary figure S12.** Sensitivity analysis 1, excluding participants with health conditions already present before index infection (cardiovascular diseases, respiratory diseases, mental disorders, neurologic or sensory disorders, cancer, metabolic diseases, n=599) and cases with an alternative explanation of persisting symptoms (n=41). Shown are cardiopulmonary function indicators and grip strength (means with 95%-CI) by case-control status at clinical examination in phase 2. Adjusted for sex-age class combinations, study centre, university entrance qualification, BMI, smoking status and use of beta blocking agents. For comparability the x-axis is scaled from mean -1 SD to mean +1 SD for all panels.

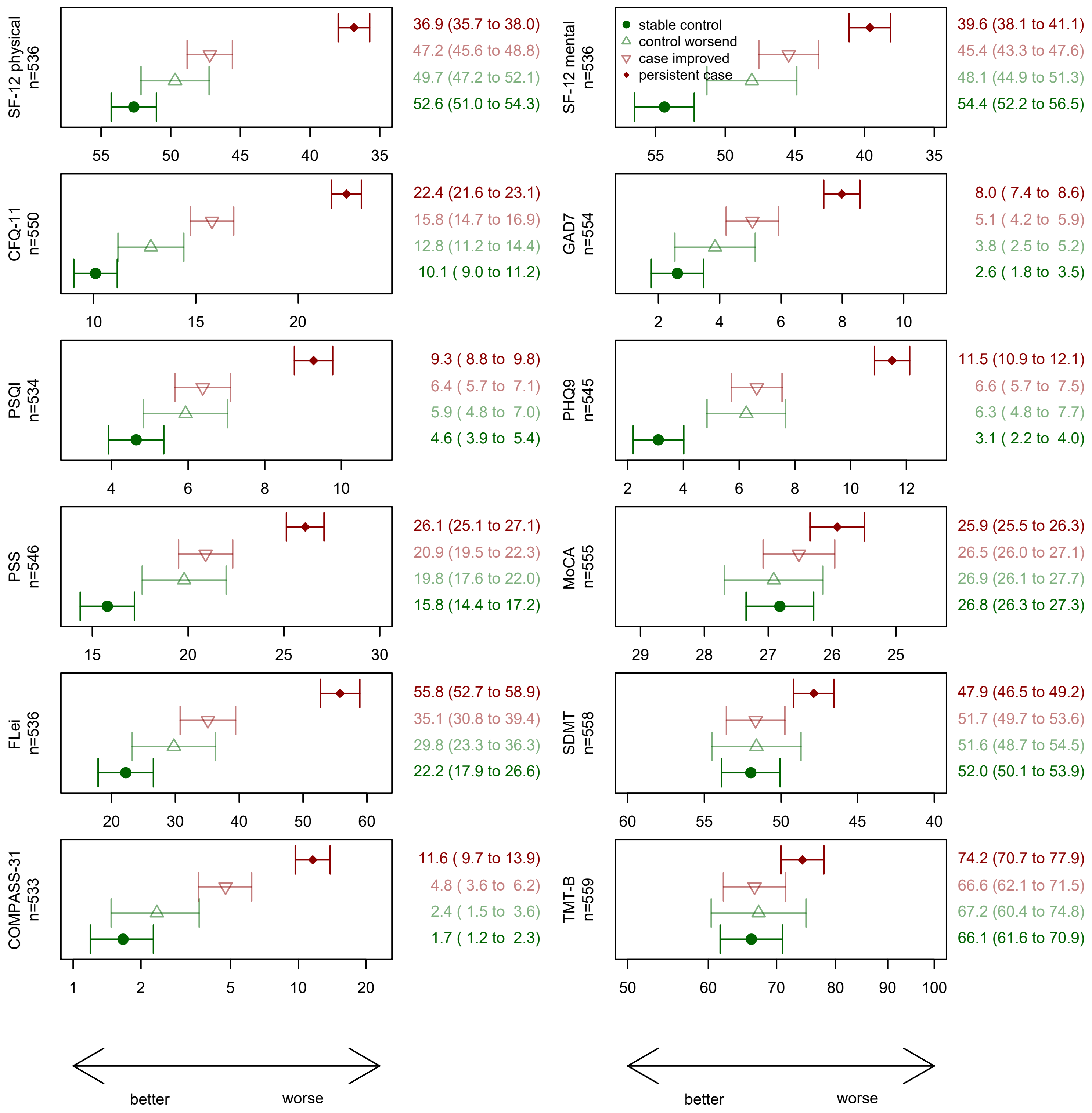

**Supplementary figure S13.** Sensitivity analysis 2, showing results for study participants with a BMI ≥27.5 kg/m². Shown are means (geometric mean for COMPASS-31 and TMT-B) of self-reported health outcomes and neurocognitive tests (with 95%-CI) by case-control status at clinical examination in phase 2, adjusted for sex-age class combinations, study centre, and university entrance qualification. For comparability the x-axis is scaled from mean -1 SD to mean +1 SD for all panels. MoCA: Montreal cognitive assessment scale (points); SDMT: Symbol Digit Modalities Test (number of correct symbols); TMT-B: Trail making test B (time in seconds).

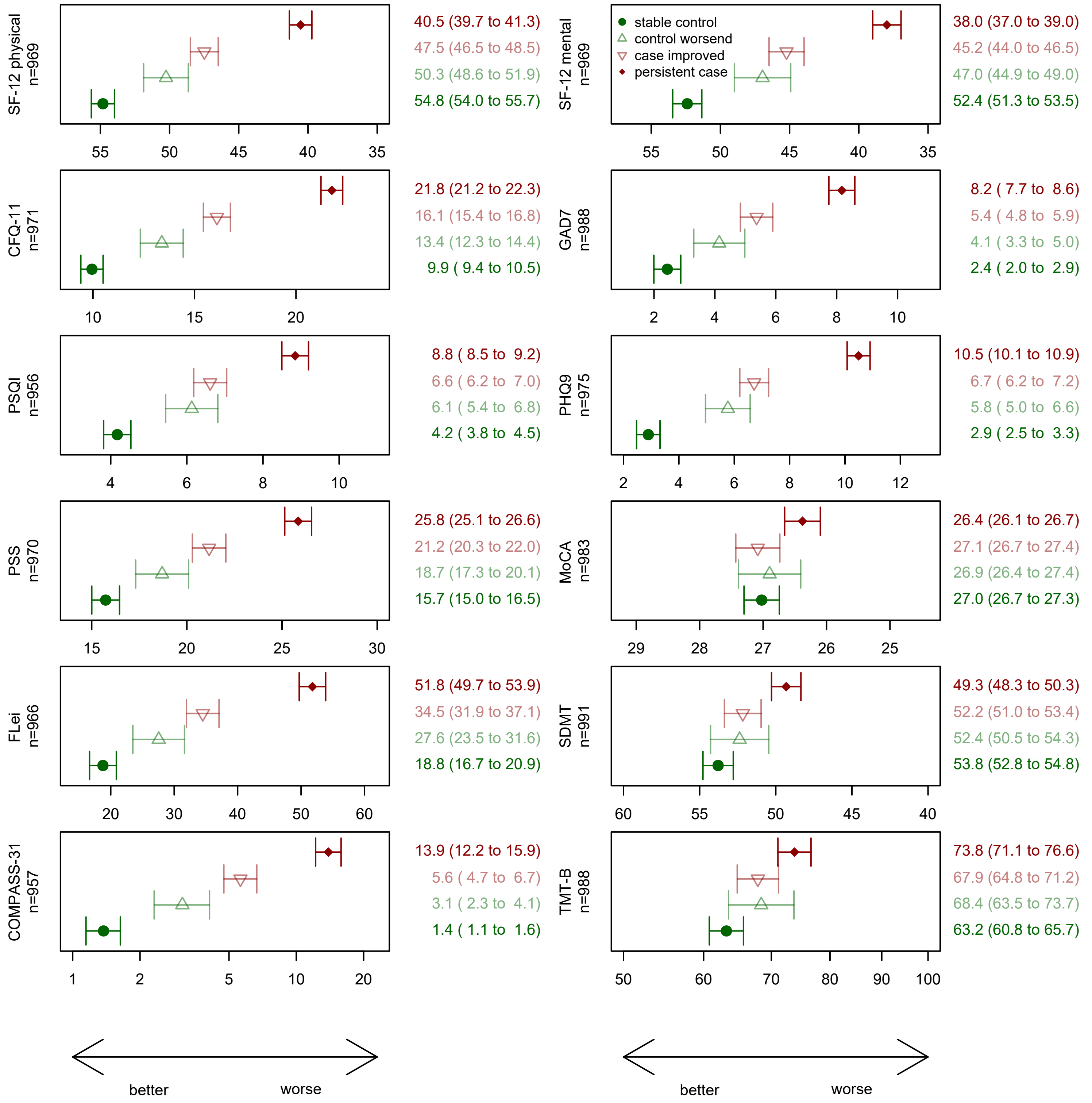

**Supplementary figure S14.** Sensitivity analysis 2, results for study participants with a BMI <27.5 kg/m². Shown are means (geometric mean for COMPASS-31 and TMT-B) of self-reported health outcomes and neurocognitive tests (with 95%-CI) by case-control status at clinical examination in phase 2, adjusted for sex-age class combinations, study centre, and university entrance qualification. For comparability the x-axis is scaled from mean -1 SD to mean +1 SD for all panels. MoCA: Montreal cognitive assessment scale (points); SDMT: Symbol Digit Modalities Test (number of correct symbols); TMT-B: Trail making test B (time in seconds).

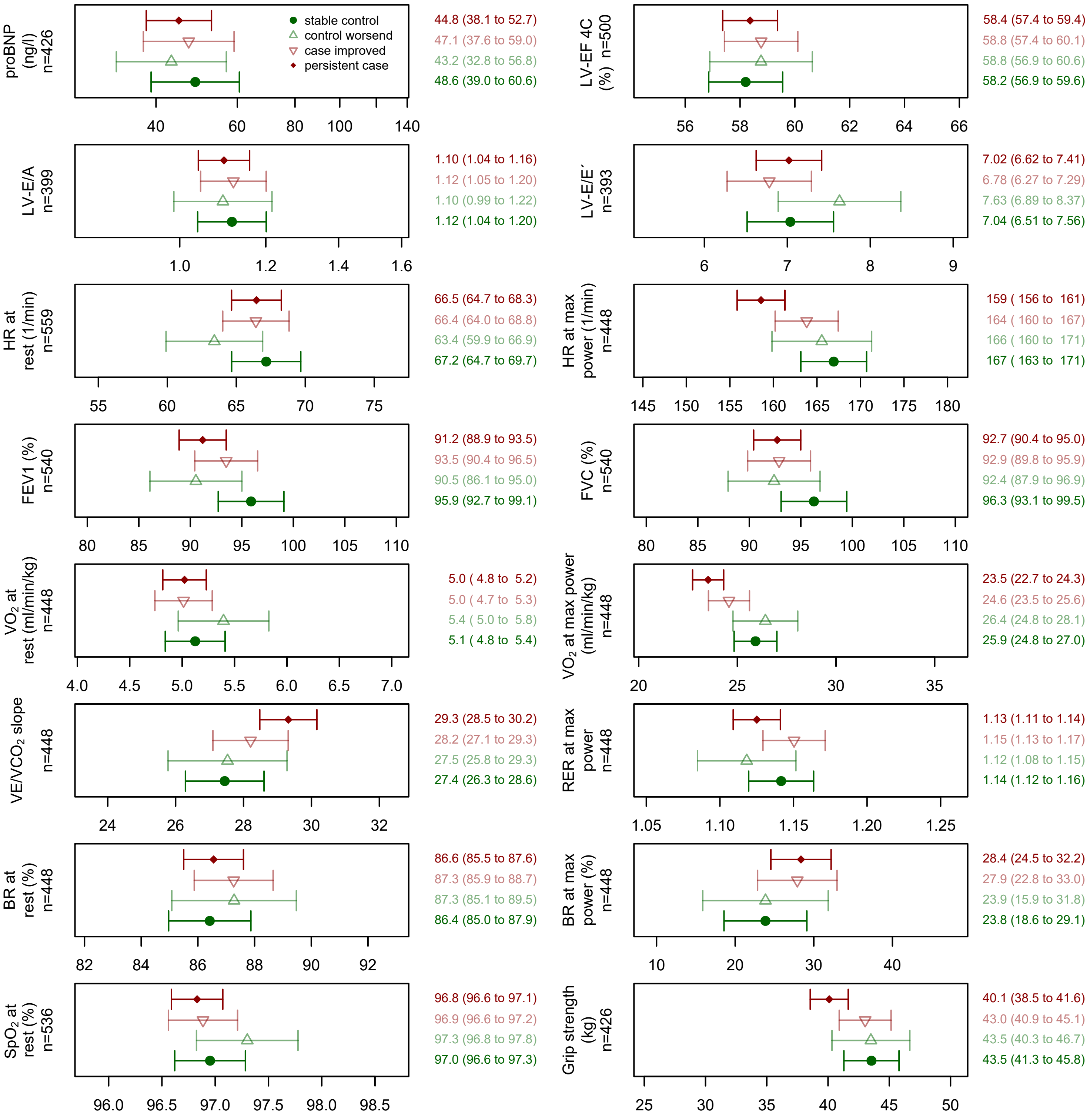

**Supplementary figure S15.** Sensitivity analysis 2, results for study participants with a BMI ≥27.5 kg/m². Shown are cardiopulmonary function indicators and grip strength (means with 95%-CI) by case-control status at clinical examination in phase 2. Adjusted for sex-age class combinations, study centre, university entrance qualification, smoking status and use of beta blocking agents. For comparability the x-axis is scaled from mean -1 SD to mean +1 SD for all panels.

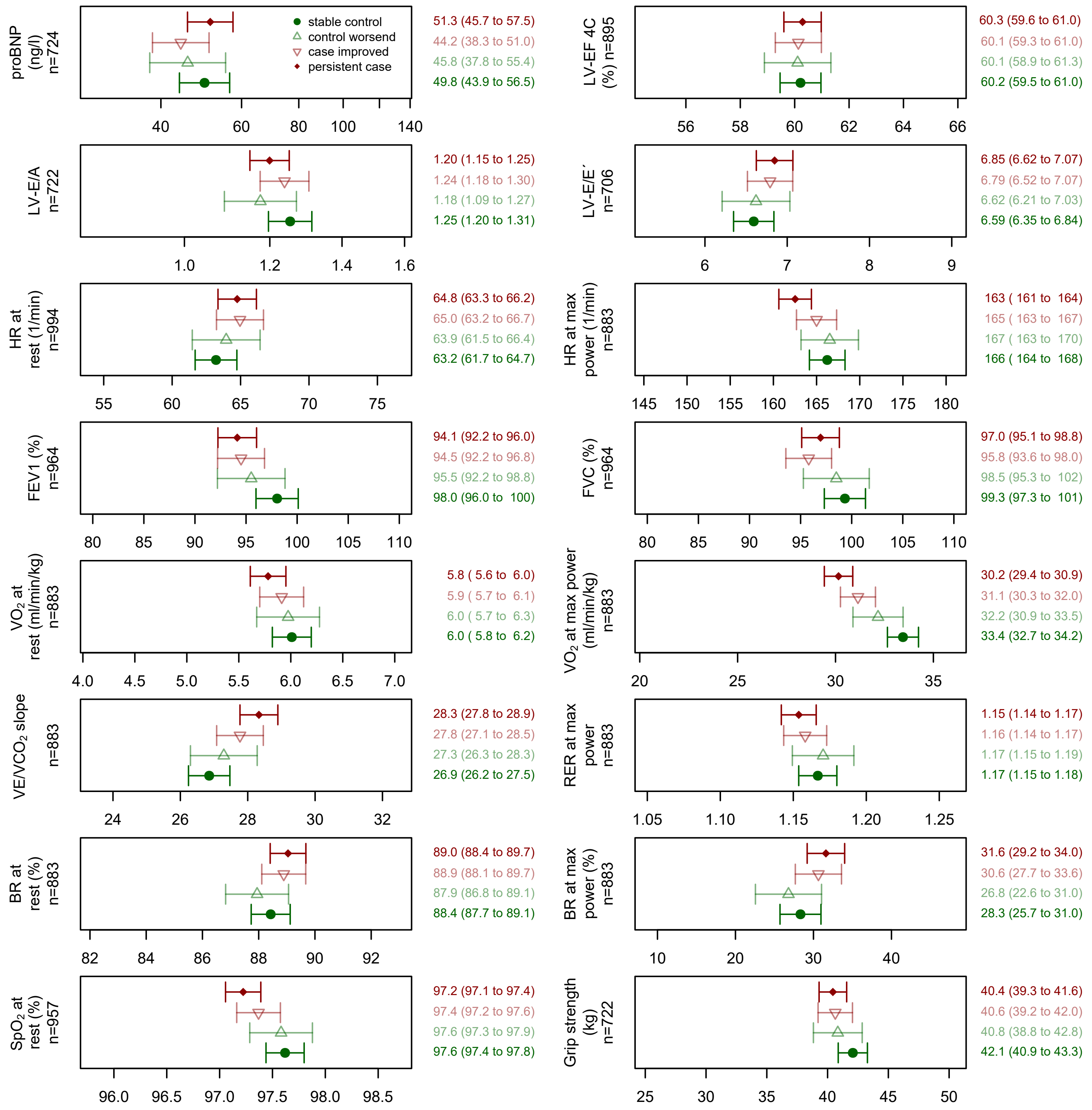

**Supplementary figure S16.** Sensitivity analysis 2, results for study participants with a BMI <27.5 kg/m². Shown are cardiopulmonary function indicators and grip strength (means with 95%-CI) by case-control status at clinical examination in phase 2. Adjusted for sex-age class combinations, study centre, university entrance qualification, smoking status and use of beta blocking agents. For comparability the x-axis is scaled from mean -1 SD to mean +1 SD for all panels.

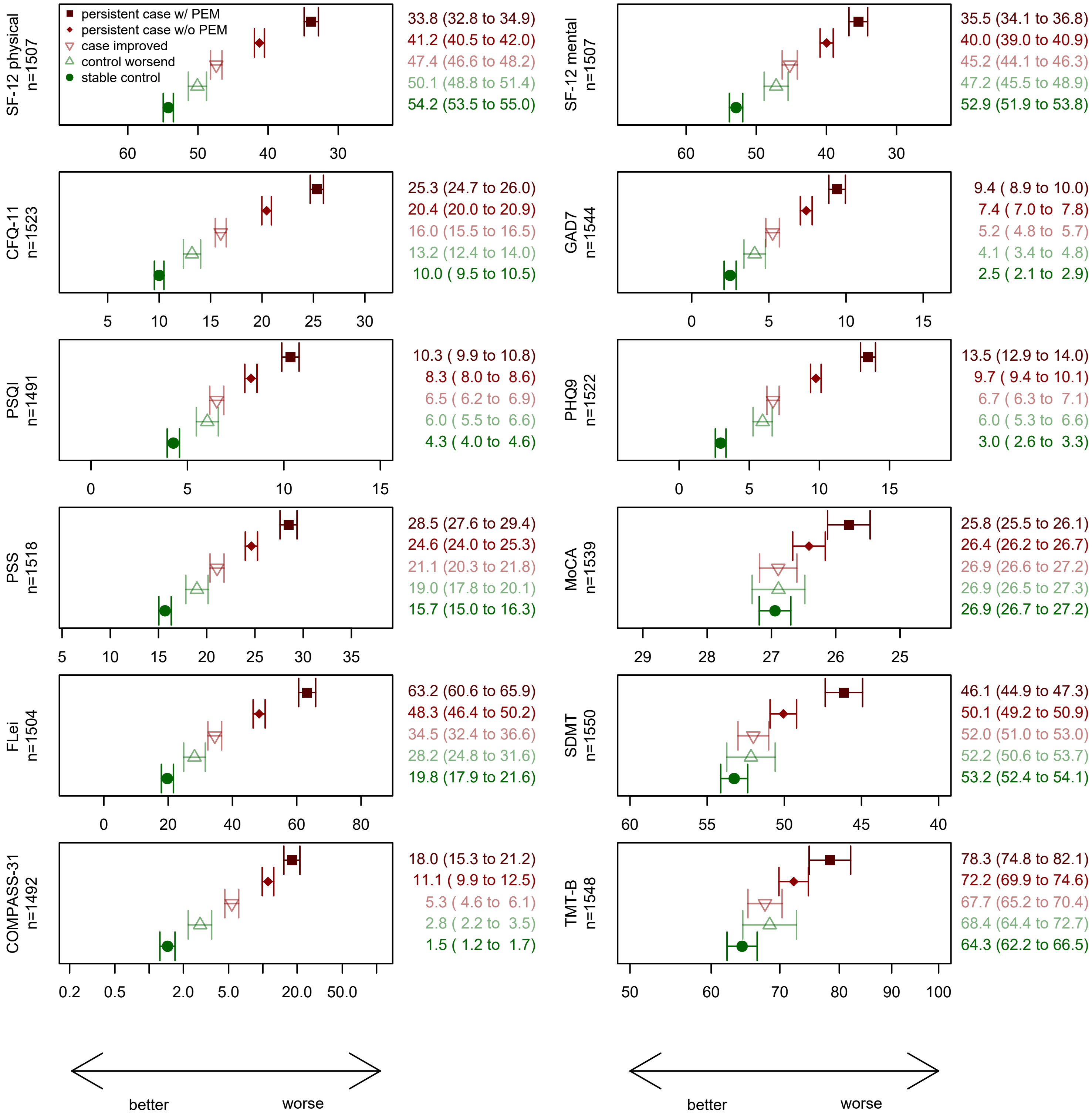

**Supplementary figure S17.** Sensitivity analysis 3 with persistent cases additionally stratified by presence of post-exertional malaise (PEM, lasting >14 hours). Shown are means (geometric mean for COMPASS-31 and TMT-B) of self-reported health outcomes and neurocognitive tests (with 95%-CI) at clinical examination in phase 2, adjusted for sex-age class combinations, study centre, and university entrance qualification. For comparability the x-axis is scaled from mean -2 SD to mean +2 SD for all panels. MoCA: Montreal cognitive assessment scale (points); SDMT: Symbol Digit Modalities Test (number of correct symbols); TMT-B: Trail making test B (time in seconds).

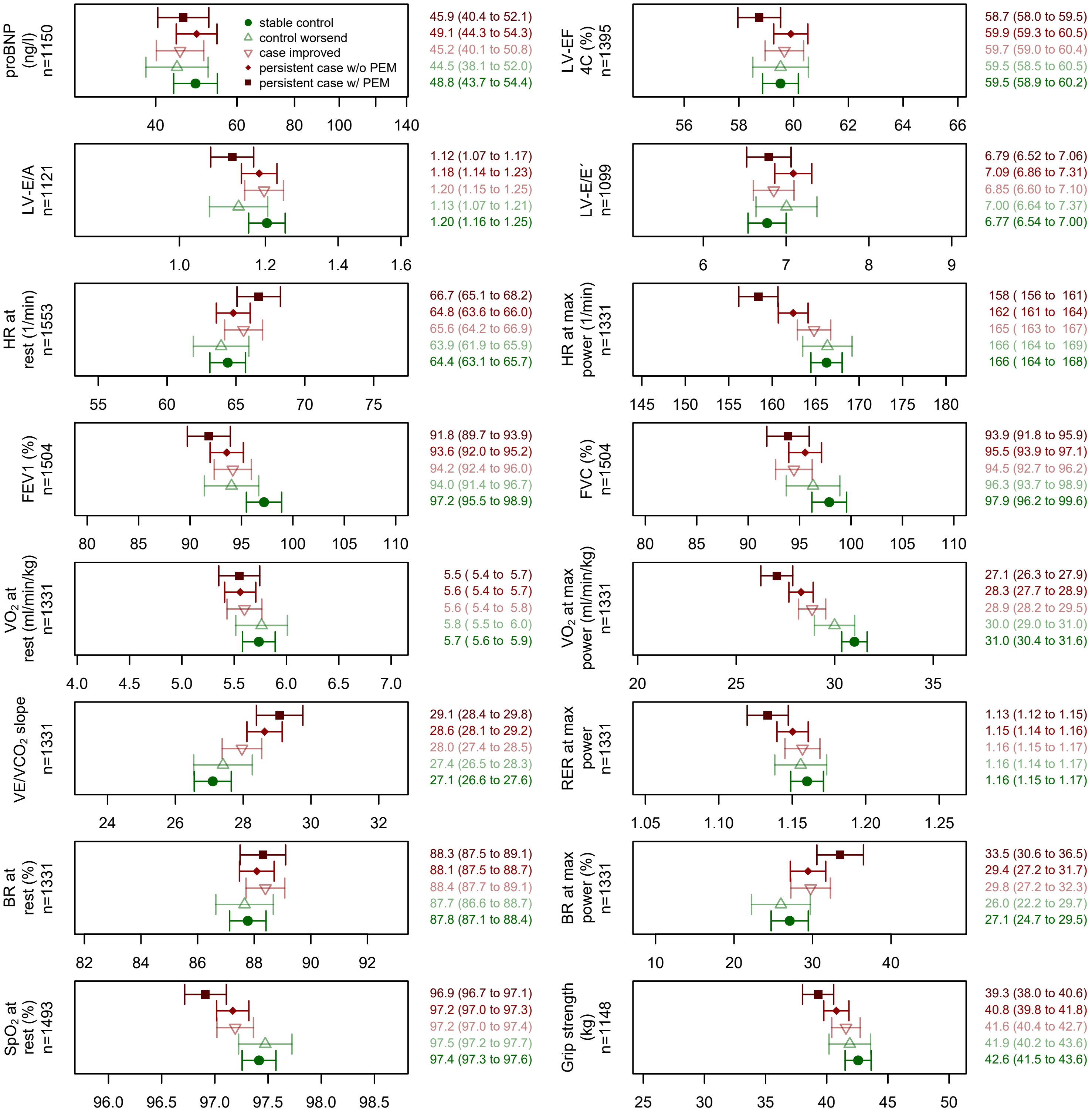 

**Supplementary figure S18.** Sensitivity analysis 3 with persistent cases additionally stratified by presence of post-exertional malaise (PEM, lasting >14 hours). Shown are cardiopulmonary function indicators and handgrip strength (means with 95%-CI) at clinical examination in phase 2. Adjusted for sex-age class combinations, study centre, university entrance qualification, BMI, smoking status and use of beta blocking agents. For comparability the x-axis is scaled from mean -1 SD to mean +1 SD for all panels.

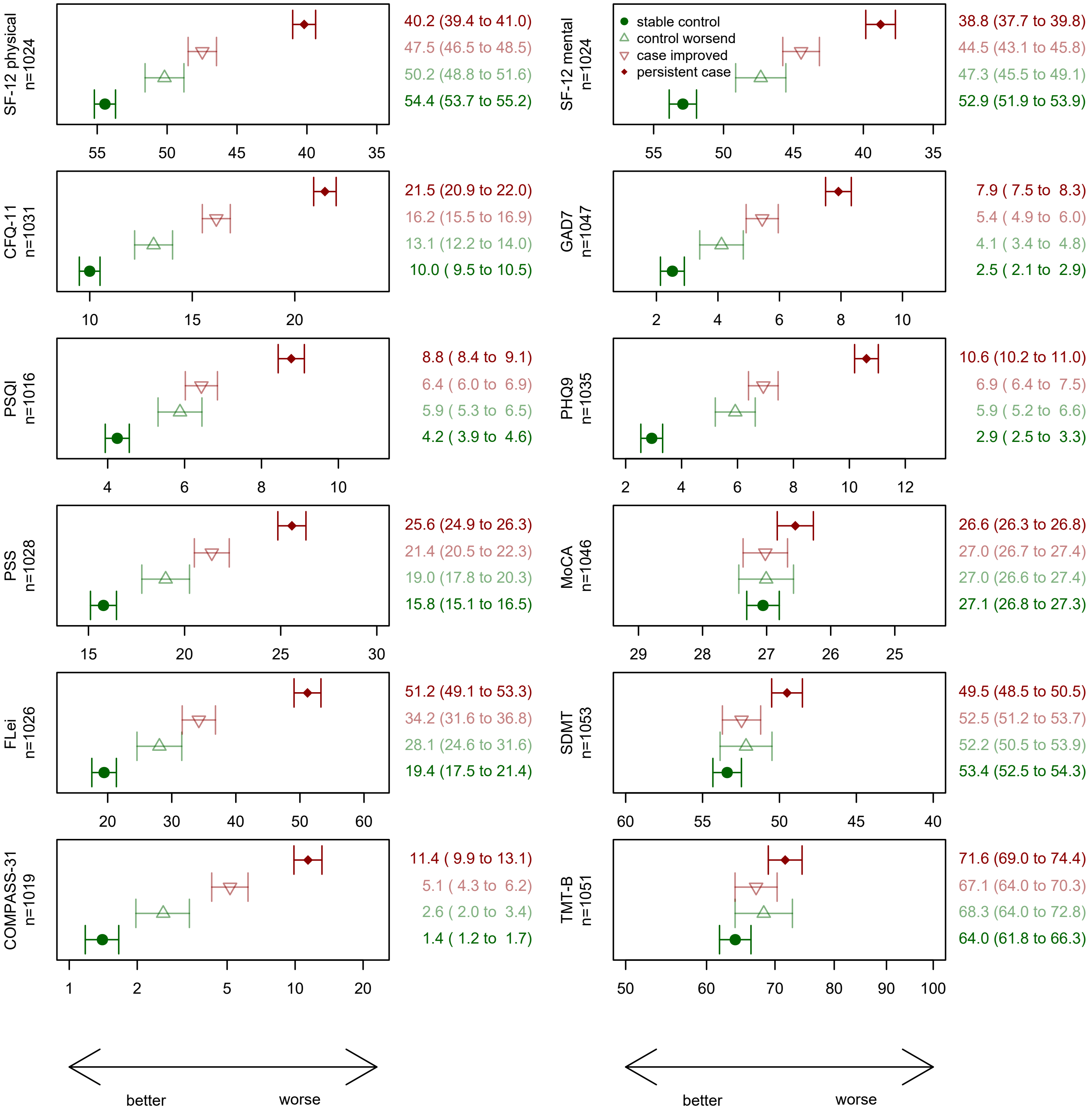

**Supplementary figure S19.** Sensitivity analysis 4, in participants without medical care for their earlier acute (index) SARS-CoV-2 infection. Shown are means (geometric mean for COMPASS-31) of self-reported health outcomes (with 95%-CI) by stable case-control status at clinical examination in phase 2, adjusted for sex-age class combinations and university entrance qualification. For comparability the x-axis is scaled from mean -1 SD to mean +1 SD for all panels.

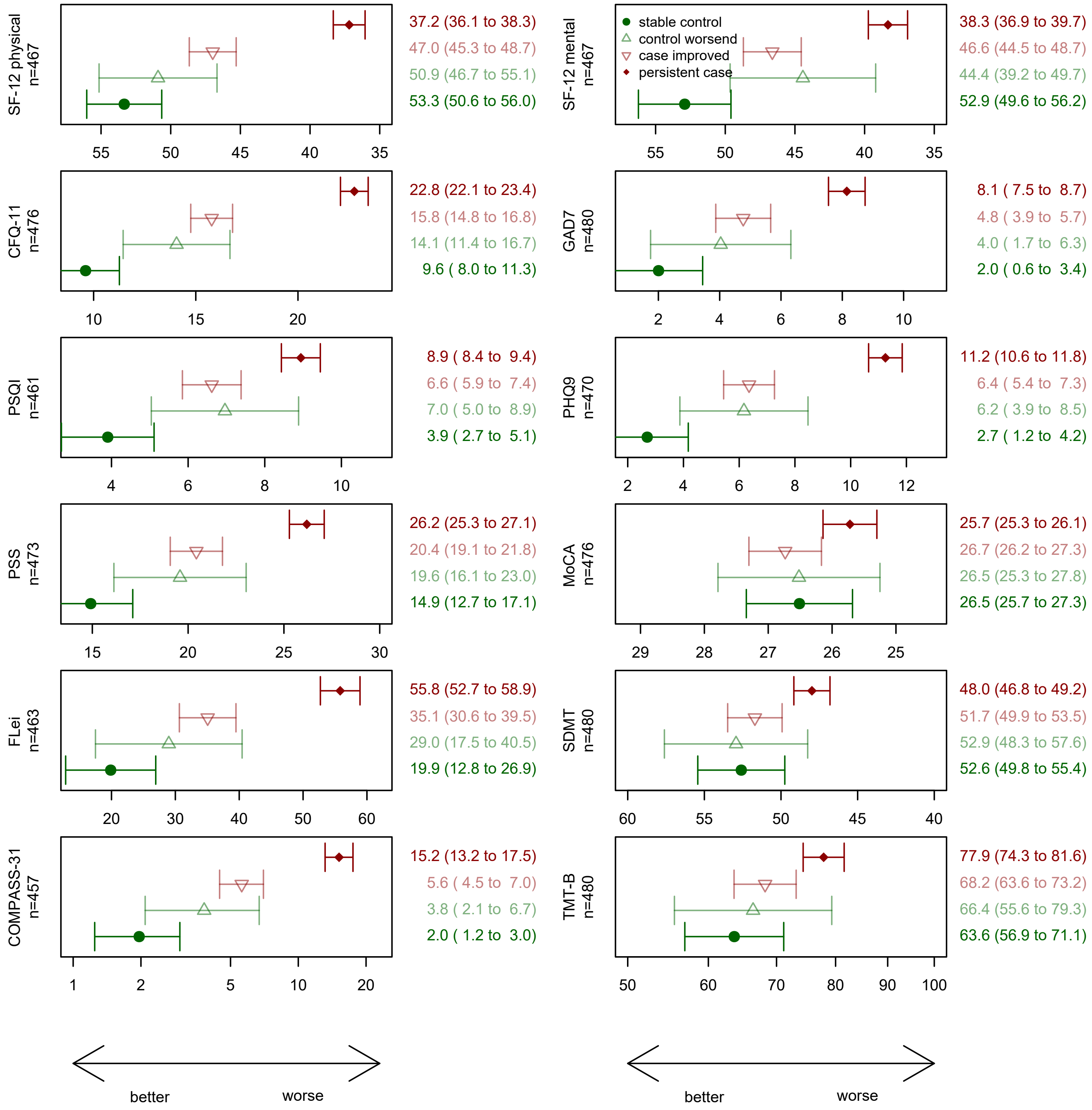

**Supplementary figure S20.** Sensitivity analysis 4, in participants with medical care for their earlier acute (index) SARS-CoV-2 infection. Shown are means (geometric mean for COMPASS-31) of self-reported health outcomes (with 95%-CI) by stable case-control status at clinical examination in phase 2, adjusted for sex-age class combinations and university entrance qualification. For comparability the x-axis is scaled from mean -1 SD to mean +1 SD for all panels.

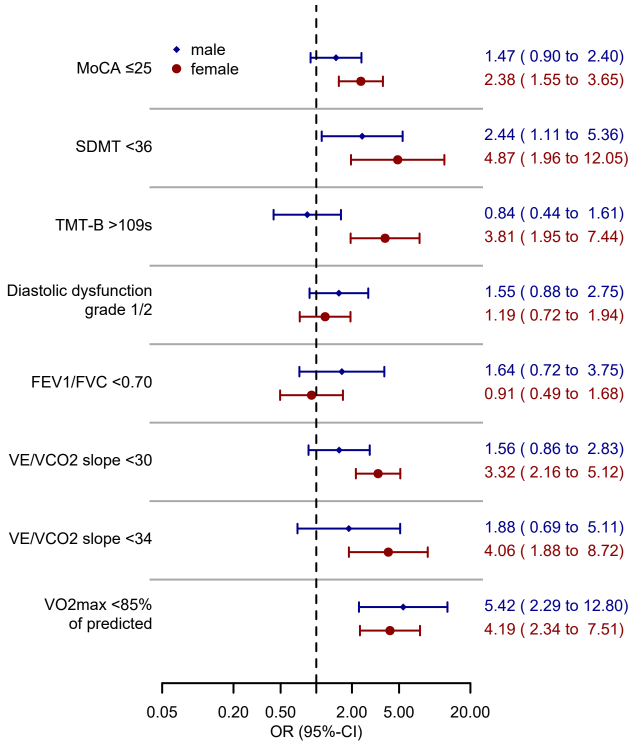

**Supplementary figure S21.** Sex specific association of case control status (persistent cases vs. stable controls) witch abnormal neurocognitive and cardiopulmonary test results.
