## Supplementary text for "Persistent symptoms and clinical findings in adults with post-acute sequelae of COVID-19/post-COVID-19 syndrome in the second year after acute infection: population-based, nested case-control study"

### Additional information about methods

**Supplementary text S1**. Details of clinical assessments and validated questionnaires (with references)

**mMRC**. The mMRC dyspnoea scale assesses the disability due to breathlessness. It comprises five grades describing different activities and moderately correlates with other healthcare-associated morbidity, mortality and quality of life scales (particularly in COPD). The instrument is commonly used and has recently been recommended for COVID-19 and long COVID research.

- Mahler DA, Wells CK. Evaluation of clinical methods for rating dyspnea. *Chest* 1988; 93:580-6.
- Gorst SL, Seylanova N, Dodd SR, Harman NL, O'Hara M, Terwee CB, et al. Core outcome measurement instruments for use in clinical and research settings for adults with post-COVID-19 condition: an international Delphi consensus study. *Lancet Respir Med* 2023; 11:1101-14.

**Handgrip strength test**. The grip strength test measures the maximum isometric strength of the hand and forearm muscles. It uses a hydraulic dynamometer with the arm positioned at right angles and the elbow by the side of the body. The test subject squeezes the dynamometer with maximum isometric effort for about 5 seconds. Both hands are grip tested three times (in an alternating manner), and the maximum strength is recorded. We used different devices, all CE marked and calibrated, including SAEHAN® DHD-1, SH5001 and SH1003 hydraulic hand dynamometer devices (Saehan Corporation, Changwon-si, South Korea).

- Roberts HC, Denison HJ, Martin HJ, Patel HP, Syddall H, Cooper C, et al. A review of the measurement of grip strength in clinical and epidemiological studies: towards a standardised approach. *Age Ageing* 2011; 40:423-9.

**Multifrequency bioelectrical impedance analysis**. Whole body composition was measured using one of the following instruments: Inbody 770 (Biospace Korea, Seoul, South Korea), BIA 101 BIVA® PRO (Akern s.r.l., Florence, Italy), InBody 4.0 (InBody Europe B.V., Eschborn, Deutschland). The percentage of body fat was measured according to standard procedures.

- Kyle UG, Bosaeus I, De Lorenzo AD, Deurenberg P, Elia M, Gómez JM, et al. Bioelectrical impedance analysis--part I: review of principles and methods. *Clin Nutr* 2004; 23:1226-43.

**Validated questionnaires – health-related quality of life**. We used the SF-12 Health Survey, a short form of the SF-36 instrument, to assess health-related quality of life. The 12 items are rated on a 5-point Likert scale and can be evaluated on two subscales: a mental component and a physical component. The German version has been tested in healthy individuals and other populations.

- Ware J, Kosinski M, Keller SD. A 12-Item Short-Form Health Survey: construction of scales and preliminary tests of reliability and validity. *Med Care* 1996; 34:220-33.
- Gandek B, Ware JE, Aaronson NK, Apolone G, Bjorner JB, Brazier JE, et al. Cross-validation of item selection and scoring for the SF-12 Health Survey in nine countries: results from the International Quality of Life Assessment project. *J Clin Epidemiol* 1998; 51:1171-8.
- Wirtz MA, Morfeld M, Glaesmer H, Brähler E. Normierung des SF-12 Version 2.0 zur Messung der gesundheitsbezogenen Lebensqualität in einer deutschen bevölkerungsrepräsentativen Stichprobe. *Diagnostica* 2018; 64:215-26.

**Validated questionnaires – fatigue and sleep**. We used the CFQ-11 to assess the extent and severity of fatigue. The instrument is available in German and has been tested in a representative sample of the German population. Each of the 11 items is answered on a 4-point Likert scale, yielding a maximal global score of 33 or a maximal binary/bimodal score of 11. A bimodal score of >3 qualifies for "caseness", and a total score of >29 indicates extreme fatigue.

- Chalder T, Berelowitz G, Pawlikowska T, Watts L, Wessely S, Wright D, et al. Development of a fatigue scale. *J Psychosom Res* 1993; 37:147-53.
- Martin A, Staufenbiel T, Gaab J, Rief W, Brähler E. Messung chronischer Erschöpfung – teststatistische Prüfung der Fatigue Skala (FS). *Z Klin Psychol Psychother* 2010; 39:33-44.

- Cella M, Chalder T. Measuring fatigue in clinical and community settings. *J Psychosom Res* 2010; 69:17-22.

The PSQI was used to evaluate overall sleep quality (over the past month). The 19 items belong to one of seven subcategories: subjective sleep quality, sleep latency, sleep duration, habitual sleep efficiency, sleep disturbances, use of sleeping medication, and daytime dysfunction. A German version has been validated in healthy individuals and other populations.

- Buysse DJ, Reynolds CF, Charles F, Monk TH, Berman SR, Kupfer DJ. The Pittsburgh sleep quality index: a new instrument for psychiatric practice and research. *Psychiatry Research* 1989; 28:193-213.

- Backhaus J, Riemann D. Schlafstörungen bewältigen. Weinheim: Beltz Psychologie Verlags Union; 1996.

The ISI is a short 7-item screening instrument to assess the nature, severity, and impact of insomnia in the past two weeks. Its German version has been validated for the evaluation of the subjective perception of sleep complaints and compared with the PSQI in healthy individuals and other populations. ISI scores range from 0 to 28 and can be interpreted as follows: <8, no insomnia, >14 insomnia (>21 severe insomnia). There was a high correlation between PSQI and ISI scores ($r=0.816$, $p<0.001$) but weaker correlations ($r<0.5$) between PSQI or ISI with ESS in the EPILOC phase 2 participant population.

- Bastien CH, Vallières A, Morin CM. Validation of the Insomnia Severity Index (ISI) as an outcome measure for insomnia research. *Sleep Med* 2001; 2:297-307.
- Morin CM, Belleville G, Bélanger L, Ivers H. The Insomnia Severity Index: psychometric indicators to detect insomnia cases and evaluate treatment response. *Sleep* 2011; 34:601-8.
- Gerber M, Lang C, Lemola S, Colledge F, Kalak N, Holsboer-Trachsler E, et al. Validation of the German version of the insomnia severity index in adolescents, young adults and adult workers: results from three cross-sectional studies. *BMC Psychiatry* 2016; 16:174.

The ESS was used to assess daytime sleepiness in middle-aged white individuals. Respondents rate their usual chances of dozing off or falling asleep in eight different daily situations on a 4-point scale. The total ESS score ranges between 0 and 24, with a higher score reflecting a higher level of daytime sleepiness. A score of <11 corresponds to the normal range of sleepiness in healthy adults. A validated German version of the ESS is available. In the EPILOC phase 2 participant population, the correlation between ESS, PSQI, and ISI was moderate.

- Kendzerska TB, Smith PM, Brignardello-Petersen R, Leung RS, Tomlinson GA. Evaluation of the measurement properties of the Epworth sleepiness scale: a systematic review. *Sleep Med Rev* 2014; 18:321-31.
- Bloch KE, Schoch OD, Zhang JN, Russi EW. German version of the Epworth Sleepiness Scale. *Respiration* 1999; 66:440-7.
- Sander C, Hegerl U, Wirkner K, Walter N, Kocalevent RD, Petrowski K, et al. Normative values of the Epworth Sleepiness Scale (ESS), derived from a large German sample. *Sleep Breath* 2016; 20:1337-45.

**Validated questionnaires – mood and anxiety**. We used the 9-item PHQ-9 to screen for depressive symptom severity and the 7-item GAD-7 to assess anxiety. The PHQ-9 has strong diagnostic accuracy in assessing depression. Respondents are asked to rate each item on a 0 to 3 Likert-type scale how frequently they experienced each symptom over the past 2 weeks (0, not at all; 3, nearly every day). Scores are summed to yield a total score ranging from 0 to 27. Cut-off scores between 8 and 11 have good sensitivity and specificity for detecting depression, and values >14 indicate moderate to severe depression. A German version is available.

- Kroenke K, Spitzer RL, Williams JBW, Löwe B. The Patient Health Questionnaire Somatic, Anxiety, and Depressive Symptom Scales: a systematic review. *Gen Hosp Psychiatry* 2010;32:345–59.
- Manea L, Gilbody S, McMillan D. Optimal cut-off score for diagnosing depression with the Patient Health Questionnaire (PHQ-9): a meta-analysis. *CMAJ* 2012; 184:E191-6.
- Gräfe K, Zipfel S, Herzog W, Löwe B. Screening psychischer Störungen mit dem "Gesundheitsfragebogen für Patienten (PHQ-D)". Ergebnisse der deutschen Validierungsstudie. *Diagnostica* 2004; 50:171-81.

The GAD-7, originally developed to screen for generalised anxiety disorders in primary care settings, is now commonly used across various settings and populations. It has also been used to screen for post-traumatic stress disorder, social anxiety or panic disorders. The total score for the seven items ranges from 0 to 21. A score >9 indicates moderate to severe anxiety. A validated German version is available.

- Spitzer RL, Kroenke K, Williams JBW, Löwe B. A brief measure for assessing generalised anxiety disorder: the GAD-7. *Arch Intern Med* 2006; 166:1092-7.
- Löwe B, Müller S, Brähler E, Kroenke K, Albani C, Decker O. Validierung und Normierung eines kurzen Selbstratinginstrumentes zur Generalisierten Angst (GAD-7) in einer repräsentativen Stichprobe der deutschen Allgemeinbevölkerung. *Psychother Psychosom Med Psychol* 2007; 57:A050.

**Validated questionnaires – perceived stress**. The PSS-10 is a 10-item questionnaire developed to assess stress levels. It evaluates the degree to which an individual has perceived life as unpredictable, uncontrollable and overloading over the previous month and captures perceived helplessness and self-efficacy. PSS-10 scores correlate with depression, anxiety and fatigue. A German version has been validated in healthy individuals and patients with diverse mental illnesses.

- Cohen S, Kamarck T, Mermelstein R. A global measure of perceived stress. *J Health Soc Behav* 1983; 24:385-96.
- Klein EM, Brähler E, Dreier M, Reinecke L, Müller KW, Schmutzer G, et al. The German version of the Perceived Stress Scale - psychometric characteristics in a representative German community sample. *BMC Psychiatry* 2016; 16:159.
- Schneider EE, Schönfelder S, Domke-Wolf M, Wessa M. Measuring stress in clinical and nonclinical subjects using a German adaptation of the Perceived Stress Scale. *Int J Clin Health Psychol* 2020; 20:173-81.

**Validated questionnaires – cognitive complaints**. FLei is a questionnaire designed to assess subjective cognitive complaints. It is a 35-item instrument using a five-point rating scale (never; rarely; sometimes; often; very often) focusing on difficulties in everyday situations in the last 6 months in three areas (attention, memory and executive functioning). The subscores range between 0 and 40, and the total score ranges between 0 and 120, with higher scores indicating lower subjective cognitive ability. The FLei has been developed primarily for patients with mental disorders. It has earlier been used in patients with PCS, for whom the mean total score was 69, and both the attention (mean, 25) and memory (mean, 26) subscores were higher than the executive subscore (mean 18). Also, there was a high correlation between the FLei total score and fatigue.

- Beblo T, Kunz M, Brokate B, Scheurich A, Weber B, Albert A, et al. Entwicklung eines Fragebogens zur subjektiven Einschätzung der geistigen Leistungsfähigkeit (FLei) bei Patienten mit psychischen Störungen. *Z Neuropsychol* 2010; 21:143-51.
- Delgado-Alonso C, Díez-Cirarda M, Pagán J, Pérez-Izquierdo C, Oliver-Mas S, Fernández-Romero L, et al. Unraveling brain fog in post-COVID syndrome: Relationship between subjective cognitive complaints and cognitive function, fatigue, and neuropsychiatric symptoms. *Eur J Neurol* 2023 Oct 5. doi: 10.1111/ene.16084.

**Validated questionnaires – dysautonomia**. We used the COMPASS-31 instrument to assess symptoms suspicious of autonomic nervous system dysfunction through six weighted symptom domains (orthostatic intolerance, vasomotor, secretomotor, gastrointestinal, bladder, and pupillomotor. The questionnaire generates a weighted score from 0 to 100, with higher scores indicating worse autonomic dysfunction. A score of >19 suggests moderate-to-severe dysautonomia. In an earlier study with Long COVID patients, moderate-to-severe dysautonomia, as measured by an increased COMPASS-31 score, was frequent (66%) but not clearly linked to functional outcomes and quality of life. It has also been shown that the correlation between the COMPASS-31 score and instruments of objective measurement of dysautonomia is rather limited. A German version of the COMPASS-31 instrument has been validated.

- Sletten DM, Suarez GA, Low PA. COMPASS 31: A refined and abbreviated composite autonomic symptom score. *Mayo Clinic Proc* 2012; 87:1196-201.
- Larsen NW, Stiles LE, Shaik R, Schneider L, Muppidi S, Tsui CT, et al. Characterisation of autonomic symptom burden in long COVID: a global survey of 2,314 adults. *Front Neurol* 2022; 13:1012668.

- Novak P, Systrom DM, Marciano SP, Knief A, Felsenstein D, Giannetti MP, et al. Mismatch between subjective and objective dysautonomia. *Sci Rep* 2024; 14:2513.
- Goldstein DS. Post-COVID dysautonomias: what we know and (mainly) what we don't know. *Nat Rev Neurol* 2024; 20:99-113.
- Hilz MJ, Wang R, Singer W. Validation of the Composite Autonomic Symptom Score 31 in the German language. *Neurol Sci* 2022; 43:365-71.

**Cognitive tests**. Cognitive status was assessed using the MoCA test, the SDMT and the TMT-B. In a subgroup of study participants (with a FLei memory subscore >19) we also performed the "Verbaler Lern- and Merkfähigkeitstest" (VLMT; Helmstaedter, Lendt & Lux: Verbaler Lern- und Merkfähigkeitstest. 1. Auflage 2001, Beltz Test, Göttingen) which is a German version of the "Auditory Verbal Learning Test" (AVLT; Lezak MD: Neuropsychological Assessment. 2nd Edition, 1983, Oxford University Press, New York). This test requires wordlist learning and allows the assessment of different memory parameters in one testing session. The results of this assessment will be analysed later and reported elsewhere.

The MoCA was validated as a sensitive tool for early detection of mild cognitive impairment. The instrument evaluates some cognitive functions, including visuospatial ability, executive function, short-term and long-term memory recall, attention, language, abstraction, delayed recall, and orientation. The maximum score is 30 points. The original validation study suggested to use a cut-off of ≤26. Subsequent studies have shown that lower cut-off values (23 or 24) may yield fewer false-positive indications of cognitive impairment.

- Nasreddine ZS, Phillips NA, Bédirian V, Charbonneau S, Whitehead V, Collin I, et al. The Montreal Cognitive Assessment, MoCA: a brief screening tool for mild cognitive impairment. *J Am Geriatr Soc* 2005; 53:695-9.
- Thomann AE, Berres M, Goettel N, Steiner LA, Monsch AU. Enhanced diagnostic accuracy for neurocognitive disorders: a revised cut-off approach for the Montreal Cognitive Assessment. *Alzheimers Res Ther* 2020; 12:39.
- Malek-Ahmadi M, Nikkhahmanesh N. Meta-analysis of Montreal cognitive assessment diagnostic accuracy in amnestic mild cognitive impairment. *Front Psychol* 2024; 15:1369766.

The SDMT evaluates attention, visual scanning, motor speed, and associative learning. The test requires participants to match 9 abstract symbols paired with numerical digits. The scores are obtained after counting the correct number of associations within 90 seconds. It ranges from 0 to 110, with higher numbers indicating better performance. The test was performed as a written test according to the SDMT Manual ($13^{th}$ version, 2013). Normal scores vary by age, gender, and education.

- Smith A. Symbol Digits Modalities Test. Western Psychological Services, Los Angeles, 1982.
- Ryan J, et al. Normative data for the symbol digit modalities test in older white Australians and Americans, African-Americans, and Hispanic/latinos. *J Alzheimer's Dis Rep* 2020; 4:313-23.
- Fellows RP, Schmitter-Edgecombe M. Symbol digit modalities test: regression based normative data and clinical utility. Arch Clin Neuropsychol 2020; 35:105-15.

The TMT-B requires subjects to connect as quickly and correctly as possible 25 circles, including numbers and letters in an ascending pattern, alternating between the numbers and letters. The amount of time required to complete the task reflects the respective score. The instrument captures mental tracking, motor speed, cognitive flexibility and selective attention. Mean scores in healthy individuals increase with age (50 s for 18-34 years; 75 s for 60-64 years) and depend on education levels.

- Reitan RM. Trail Making Test. Manual for administration and scoring. Tucson, AZ: Reitan Neuropsychology Laboratory, 1992.
- Gaudino EA, Geisler MW, Squires NK. Construct validity in the Trail Making Test: what makes Part B harder? *J Clin Exp Neuropsychol* 1995; 17:529-35.
- Tombaugh TN. Trail Making Test A and B: normative data stratified by age and education. *Arch Clin Neuropsychol* 2004; 19:203-14.
- Bowie CR, Harvey PD. Administration and interpretation of the trail making test. *Nat Protoc* 2006; 1:2277-81.

#### **Supplementary text S2.** Methodological details of echocardiography and CPET (with references)

**Echocardiography**. Resting echocardiograms were performed with one of the following instruments: GE Vivid E9 (GE Healthcare, Solingen, Germany), EPIC7 (Philips Healthcare, Andovar, Massachusetts/USA), iE33 (Philips Medical Systems B.V., Eindhoven, Netherlands). Data sets were digitally stored and exported on a dedicated workstation. The assessment of biventricular and atrial size, LV diastolic function, LV and RV systolic function with evaluation of the LV-EF, LV-E/e' and LV-E/A was performed according to current guidelines. Images were uploaded and analysed centrally by two independent, experienced examiners on TomTec Image Arena Version IA4.6.4, TTA2.20.01 (TomTec Imaging Systems GmbH, Unterschleißheim, Germany) as imaging analysis software.

- Lang RM, Badano LP, Mor-Avi V, Afilalo J, Armstrong A, Ernande L, et al. Recommendations for cardiac chamber quantification by echocardiography in adults: an update from the American Society of Echocardiography and the European Association of Cardiovascular Imaging. *Eur Heart J Cardiovasc Imaging* 2015; 16:233-70.
- Nagueh SF, Smiseth OA, Appleton CP, Byrd BF, Dokainish H, Edvardsen T, et al. Recommendations for the evaluation of left ventricular diastolic function by echocardiography: an update from the American Society of Echocardiography and the European Association of Cardiovascular Imaging. *Eur Heart J Cardiovasc Imaging* 2016; 17:1321-60.
- Gluckman TJ, Bhave NM, Allen LA, Chung EH, Spatz ES, Ammirati E, et al. 2022 ACC Expert Consensus Decision Pathway on cardiovascular sequelae of COVID-19 in adults: myocarditis and other myocardial involvement, post-acute sequelae of SARS-CoV-2 infection, and return to play: a report of the American College of Cardiology Solution Set Oversight Committee. J Am Coll Cardiol 2022; 79:1717-56.
- Wilson MG, Hull JH, Rogers J, Pollock N, Dodd M, Haines J, et al. Cardiorespiratory considerations for return-to-play in elite athletes after COVID-19 infection: a practical guide for sport and exercise medicine physicians. Br J Sports Med 2020; 54:1157-61.
- Szabó L, Juhász V, Dohy Z, Fogarasi C, Kovács A, Lakatos BK, et al. Is cardiac involvement prevalent in highly trained athletes after SARS-CoV-2 infection? A cardiac magnetic resonance study using sex-matched and age-matched controls. Br J Sports Med 2022; 56:553-60.

**CPET**. All CPETs were performed by means of stationary ergospirometers (Quark CPET, Rome, Italy; MetaLyzer, CORTEX Biophysics, Leipzig, Germany; or Ergostik, Geratherm, Geratal, Germany) using an electronically braked cycle ergometer. Patients received a standard explanation before the procedure and wore a non-rebreathing Hans-Rudolph mask connected to the respective ergospirometry system. CPET started with a 2 min resting phase followed by an unloaded pedalling phase of 2 min, a linear ramp increase in load tailored for each patient (continuous increase equivalent to an increase of 10, 15, 20, 25, 30, 35 or 40 W per minute) in order to achieve exhaustion in 6-12 min and a recovery phase of 3 min. The tests were done according to current recommendations published by the German Respiratory Society.

During tests, pulmonary ventilation (VE), $O_2$ consumption ($VO_2$) and $CO_2$ output ($VCO_2$) were continuously measured breath-by-breath. Electrocardiogram (CardioPart 12, Amedtec, Aue, Germany; Custo med GmbH, Ottobrunn, Germany) and finger pulse oximetry (SpO2, Radical-7, Masimo, CA, USA; EDAN pulse oximeter H100B, Edan, San Diego, CA, USA) were continuously recorded. Blood pressure was measured manually. The level of dyspnoea and muscular fatigue was assessed at peak exercise using the Modified BORG Dyspnea Scale (Borg CR10), and the reason for exercise termination was recorded (fatigue, dyspnoea or undetermined). For objective determination of maximal effort, a respiratory exchange ratio (RER, $VCO_2/VO_2$) of >1.05 had been predetermined. $VO_{2max}$ was determined as an average of the last 30 s period of exercise; percent of predicted for $VO_{2max}$ was calculated according to Cooper and Storer, using the following formulas: male 50.02 – (0.384 X age) ml/kg/min; female 42.83 – (0.371 X age) ml/kg/min. Breathing reserve (BR) was continuously calculated as VE - (FEV1 x 40). The $VE/VCO_2$ slope was calculated over the linear component of

VE versus $VCO_2$. Values from 23 to 28 (males) and 26 to 30 (females), respectively, have been reported for normal healthy non-elderly adults.

- Meyer FJ, Borst MM, Buschmann HC, Claussen M, Dumitrescu D, Ewert R, et al. Belastungsuntersuchungen in der Pneumologie – Empfehlungen der Deutschen Gesellschaft für Pneumologie und Beatmungsmedizin e. V. *Pneumologie* 2018; 72:687-731.
- Borg G. Psychophysical scaling with applications in physical work and the perception of exercise. *Scand J Work Environ Health* 1990; 16 (Suppl 1): 55-8.
- Cooper B, Storer W. Exercise testing and interpretation. Cambridge University Press, Cambridge 2001.
- Mezzani A. Cardiopulmonary exercise testing: basics of methodology and measurements. Ann Am Thorac Soc 2017; 14(Suppl 1):S3-11.

#### **Supplementary text S3.** Details of laboratory investigations

**Biospecimen sampling and storage**. On the day of the outpatient assessment and prior to further study procedures, we collected venous blood (a total of 88 mL), saliva and urine from non-fasting participants. Samples were immediately analysed (see below) or stored below -20°C until further processing.

**Blood counts and clinical chemistry**. In brief, analyses included complete blood counts HbA1c (%), sodium (mmol/l), potassium (mmol/l), magnesium (mmol/l), creatinine (mg/dl), urea (mg/dl), estimated GFR (MDR and CKD-EPI G) (ml/min/1,73qm), LDH (U/l), CK (U/l), myoglobin (ng/ml), GPT/ALAT (U/l), GOT/ASAT (U/L), alcaline phosphatase (U/l), bilirubin (mg/dl), C-reactive protein or high sensitivity C-reactive protein (mg/l), TSH (mU/l), cortisol (nmol/l), ACTH (pg/ml), DHEA-S (µmol/l), Zn (µg/dl), Nt-proBNP (ng/l), ferritin (ng/ml), sTFR (mg/l), transferrin (g/l), IgE (mg/dl), 25-OH-vitamin D3 (ng/ml), CH50 (%), D-dimers (mg/l FEU), vWF antigen (%), vWF collagen binding activity (%), FVIII activity (%), urinary albumin, urinary creatinine. The assays were done at each site except for cortisol, ACTH and DHEA-S, which were measured centrally using ECLIA tests on a Cobas (Roche) instrument, and CH50, measured only for study participants in Freiburg and Heidelberg.

Different instruments and kits (such as Immulite, Innovance, Cobas, Atellica, Advia) were used for some of the measurements depending on the analyser systems available on site. All analyses were performed on CE-marked calibrated instruments following the manufacturer's instructions. Reference values were obtained from each site for each analyte, and all results were analysed with adjustments for centre. There were no significant differences between persistent cases and stable controls in the analytes not shown in the main text.

**SARS-CoV-2 serology**. Blood samples were sent to the respective diagnostic laboratories for serological analyses, and serum was prepared on the same day. Samples were either immediately analysed or stored at -20°C until further processing.

Antibodies against SARS-CoV-2 S1 receptor binding domain of the viral spike glycoprotein and nucleocapsid (N) were analysed locally. Antibodies against the SARS-CoV-2 N protein were measured using the Elecsys® Anti-SARS-CoV-2 IgG/IgM ECLIA test kit (Roche Diagnostics) using a Roche Cobas e610 or e411 module (Heidelberg, Tübingen and Ulm) or by recomWell SARS-CoV-2 IgG ELISA (Mikrogen Diagnostik) run on a Siemens BEP III analyser (Freiburg). Samples were analysed by CLIA for IgG reactive to S1 with sCOVG ELISA Assays (Siemens Healthineers) using a Siemens ADVIA centaur instrument (Freiburg, Heidelberg, Tübingen) or with the Elecsys® Anti-SARS-CoV-2 S test kit (Roche Diagnostics) on a Roche Cobas e402 instrument (Ulm). Data from FR, TÜ and UL were reported qualitatively. For samples collected in HD, diluted sera were analysed to obtain quantitative values (BAU/ml). All analyses were performed following the manufacturer's instructions.

**EBV and CMV serology**. Quantitative measurements of herpes virus-specific antibodies were performed centrally in HD. Samples were frozen and shipped for analysis. Antibodies against Epstein-Barr-Virus antigens (IgG and IgM against VCA, IgG against EBNA, IgG against EA-D) were quantified using Enzyme-Linked Immunosorbent Assays (ELISA) (Euroimmun, Lübeck, Germany; EI 2793-9601 G, EI 2795 G) using an Euroimmune Analyzer II. IgG and IgM antibodies against Cytomegalovirus (CMV) were measured by ELISA (Euroimmun; EI 2570-9601 G) on an Euroimmune Analyzer II. ELISA values <16 relative units/ml (RE/ml) were classified as negative, 16-<22 as borderline and ≥22 as positive. Samples with an IgM ratio ≥ 1.1 were classified as positive, ≥ 0.8 – 1.1 as borderline and < 0.8 as negative. Borderline and positive IgM antibodies against CMV were confirmed by immunoblot using the recomLine CMV-IgM assay (Mikrogen Diagnostik, Neuried, Germany) and an AutoBlot 3000 analyser (MedTec Biolab Inc., Durham; US). All measurements were performed and assessed according to the manufacturer's instructions. Due to few cases with IgM antibodies (EBV-VCA, 2 study participants; CMV, 11 study participants), these results were not further evaluated for differences between groups.

**SARS-CoV-2 antigen measurements in plasma**. Plasma samples from a subset of the cohort (100 persistent cases and 100 patients recovered control subjects) were analysed for traces of spike antigen using an ultrasensitive antigen ECL assay (S-PLEX SARS-CoV2 spike kit, K150ADJS), Mesoscale Discoveries, USA) on a Meso QuickPlex Q 60MM instrument, according to the manufacturer's instructions. The sensitivity of this method given by the manufacturer was 95 fg/ml; the dynamic range in our measurements was ~1-1000 pg/ml.

**SARS-CoV-2 RT-PCR in faecal samples**. Stool samples were collected by the patients at home in faecal sample collection media (9 ml of DNA-stabilisation solution R1100-250, Zymo® DNA/RNA shield faecal collection tube R1101-E, Zymo research corporation; sample stability of four weeks at room temperature ensured by the manufacturer) shortly before their appointment for the clinical examination. Stool samples provided by the participants (from Heidelberg) were directly processed for RT-PCR analysis to include samples from 156 persistent cases and 103 stable controls (that were all negative).

RNA extraction from stool samples was carried out by a semi-automatical spin-column preparation using QIAamp Viral RNA Mini Kit (Qiagen®, Germany) and Qiagen® Qiacube according to the manufacturer's instructions. Isolated RNA was stored at -80°C.140 µL of purified RNA were used as template for real-time RT-PCR performed on a (Light cycler 480 II; Roche) using the LightMix® Modular SARS-CE assay (#50-0776-96, TIB MolBiol, Berlin, Germany). Samples were classified as positive if the cycle threshold (Ct) value was ≤ 40. The housekeeping gene ß-actin served as an internal control to validate RNA isolation and reverse transcription efficiency.
